# Tuberculosis infection and hypertension: Prevalence estimates from the US National Health and Nutrition Examination Survey

**DOI:** 10.1101/2023.05.12.23289899

**Authors:** Argita D. Salindri, Sara C. Auld, Unjali P. Gujral, Elaine M. Urbina, Jason R. Andrews, Moises A. Huaman, Matthew J. Magee

## Abstract

**Objectives:** Latent Tuberculosis infection (LTBI) is marked by dynamic host-pathogen interactions with persistent low-grade inflammation and is associated with increased risk of cardiovascular diseases (CVD) including acute coronary syndrome, myocardial infarction, and stroke. However, few studies assess the relationship between LTBI and hypertension, an intermediate of CVD. We sought to determine the association between LTBI and hypertension using data representative of the adult US population.

**Methods:** We performed cross-sectional analyses using data from the 2011–2012 US National Health and Nutrition Examination Survey (NHANES). Eligible participants included adults with valid QuantiFERON-TB Gold In-Tube (QFT-GIT) test results who also had blood pressure measures and no history of TB disease. LTBI was defined by a positive QFT-GIT. We defined hypertension by either elevated measured blood pressure levels (i.e., systolic ≥130mmHg or diastolic ≥80mmHg) or known hypertension indications (i.e., self-reported previous diagnosis or use of antihypertensive medications). Analyses were performed using robust quasi-Poisson regressions and accounted for the stratified probability sampling design of NHANES.

**Results:** The overall prevalence of LTBI was 5.7% (95%CI 4.7–6.7) and hypertension was present among 48.9% (95%CI 45.2–52.7) of participants. The prevalence of hypertension was higher among those with LTBI (58.5%, 95%CI 52.4–64.5) than those without LTBI (48.3%, 95%CI 44.5–52.1) (prevalence ratio [PR]=1.2, 95%CI 1.1–1.3). However, after adjusting for confounders, the prevalence of hypertension was similar for those with and without LTBI (adjusted PR=1.0, 95%CI 0.9 –1.1). Among individuals without CVD risk factors of elevated BMI (PR_normal BMI_=1.6, 95%CI 1.2–2.0), hyperglycemia (PR_euglycemia_=1.3, 95%CI 1.1–1.5), or cigarette smoking (PR_non-smokers_=1.2, 95%CI 1.1–1.4), the unadjusted prevalence of hypertension was higher among those with LTBI vs. no LTBI.

**Conclusions:** More than half of adults with LTBI in the US had hypertension. Importantly, we observed a relationship between LTBI and hypertension among those without established CVD risk factors.

**Strengths and limitations:** *Strengths:*

- These analyses were conducted using data representative of civilian, non-institutionalized US adults, and thus, provide a robust population estimate of the prevalence of latent tuberculosis infection and hypertension in the US
- Comprehensive definitions and different cut-offs of hypertension were used (i.e., measured blood pressure level, previous diagnosis hypertension by healthcare providers) to model the association between latent tuberculosis infection and hypertension

*Limitations:*

Our findings may not be representative to other regions with higher burdens of tuberculosis
- The cross-sectional study design of NHANES prevented us from assessing the temporal relationship between latent tuberculosis infection and hypertension

**Summary:** The prevalence of hypertension was high (59%) among adults with tuberculosis infection in the U.S. In addition, we found that the prevalence of hypertension was significantly higher among adults with positive QFT without established hypertension risk factors.

## INTRODUCTION

About one-quarter of the world’s population (∼2 billion) has been infected to *Mycobacterium tuberculosis* (*Mtb*). ^1^ Among individuals infected with the bacteria, 5-10% are at risk of developing TB disease at some point in their life. ^2 3^ Tuberculosis infection (TBI), or most commonly known as latent tuberculosis infection or LTBI, is increasingly recognized as a heterogenous clinical state in which some individuals have dynamic host-pathogen interactions with persistent low-grade inflammation. This immune dysregulation has been associated with an increased risk of cardiovascular diseases (CVD) including acute coronary syndromes, myocardial infarction, and stroke. ^1 4-12^ This convergence of TBI and CVD risk poses a particular challenge for low- and middle-income countries where TBI is most prevalent and incidence of chronic non-communicable diseases, including CVD, is increasing rapidly. ^13 14^ Improved understanding of the impact of TBI on CVD risk is vital in settings where TBI and CVD are highly co-prevalent in order to design public health intervention programs aiming to reduce the burden of two diseases.

Epidemiologic data from observational cohort studies support an increased risk of CVD among people with TB disease. ^8-12^ Several studies also indicated that hypertension, an established intermediate of CVD, may be more common among patients with TB disease compared to non-TB controls ^8 11 15-17^. Furthermore, CVD was the leading contributor to post-TB mortality, accounting for 15 – 26% of deaths among TB survivors in a recent systematic review and meta-analysis. ^18^ In addition to these associations between TB disease and CVD, recent observational studies have found an association between TBI and various CVDs including acute myocardial infarction and coronary artery disease. ^9 19 20^. However, studies assessing the association between TBI and hypertension remain limited.

To date, few studies have evaluated the relationship between TBI and hypertension.

One cohort study from a large metropolitan healthcare system in the U.S. reported that individuals with TBI had greater incidence of hypertension compared to those without TBI and that rates were highest among those untreated for TBI. ^5^ Furthermore, it is unknown whether the quantitative measures of IGRA, which may indicate the underlying mycobaterial burden and has been associated with increased risks of progression to TB disease ^21-24^, is associated with hypertension. Improved understanding of the association between TBI, quantitative measures of IGRA, and and hypertension may clarify the role that TB prevention efforts in reducing the burden of CVD, both in the U.S. and globally.

Given existing knowledge gaps, we aimed to estimate the association between TBI and hypertension prevalence. We also investigated whether the magnitude of host immune responses to *Mtb* was associated with hypertension among those with positive IGRA test results.

## METHODS

### Study Design and Eligible Participants

We performed an analysis of cross-sectional data from the 2011 – 2012 US National Health and Nutrition Examination Survey (NHANES), the most recent NHANES cycle released that includes measures of TBI. NHANES is a study led by the US Centers for Disease Control and Prevention (CDC) which aims to assess the health and nutritional status of non-institutionalized civilians representative of the US population. NHANES collects demographic and health information using questionnaires administered by trained interviewers and standardized physical examinations performed in mobile examination centers. Eligible NHANES participants for our analyses were adults (≥18 years) with valid TBI test results and blood pressure measurements, and no history of TB disease (Figure 1).

**Figure 1.**
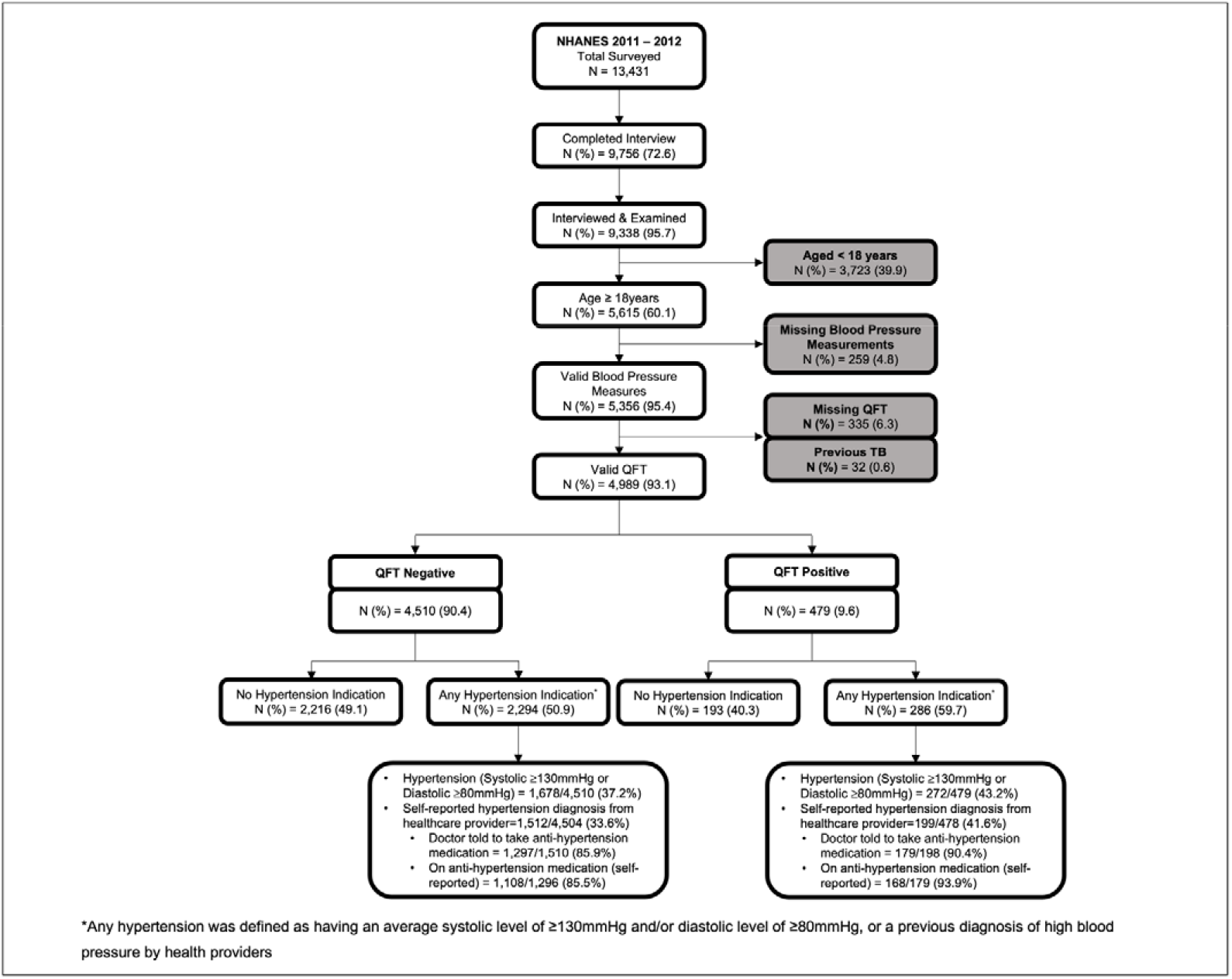
Flow chart depicting unweighted frequencies and percentages of participants included in the final analyses based on the eligibility criteria, NHANES 2011 – 2012 *This study flow chart provides description of the stepwise exclusion of ineligible participants. From 9,338 individuals who completed NHANES interview and medical examination, we included 4,989 (53*.*4%) individuals in our primary analyses after excluding those who are <18 years old or those with a record of previous TB disease, or missing blood pressure data and QuantiFERON results*

### Study Measures and Definitions

Our primary study outcome, any hypertension, was defined as having either (1) “measured hypertension,” defined as an average systolic blood pressure level of ≥130 mmHg or diastolic blood pressure level of ≥80 mmHg across three consecutive measurements, or (2) a self-reported previous hypertension diagnosis by a health care provider or current use of antihypertensive medications (i.e., known hypertension). We categorized measured blood pressure levels into “normal” (i.e., systolic <120mmHg and diastolic <80mmHg), “borderline hypertension” (i.e., systolic 120-129mmHg and diastolic <80mmHg), “stage 1 hypertension” (i.e., systolic 130 – 139mmHg or diastolic 80-89mmHg), and “stage 2 hypertension” (i.e., systolic ≥140mmHg or diastolic ≥90mmHg) according to American College of Cardiology/American Heart Association guidelines. ^25^ Among participants with a prior diagnosis of hypertension, we classified blood pressure as “controlled” (systolic <130 mmHg and diastolic <80 mmHg) or “uncontrolled” (systolic ≥130mmHg or diastolic ≥80mmHg) with or without a self-reported use of antihypertensive medications.

Our primary study exposure, TBI, was defined by a positive QuantiFERON-TB Gold In Tube or QFT test. Individuals with indeterminate test results were excluded from our analyses. For those with a positive QFT, we also extracted the quantitative results and defined the IFN-γ TB antigen response by subtracting TB NIL control values from TB antigen values (i.e., Ag-NIL values). To express IFN-γ TB antigen responses, instead of using the traditional manufacturer cut-off of ≥0.35, we used the 4.00 cut-off as previous studies showed that individuals with Ag-NIL values ≥4.00 are at greater risk from developing TB disease. ^21 23 24^ Thus, in our analyses, Ag-NIL values were categorized as “low” (<4 IU/ml) or “high” (≥4 IU/ml). For a sensitivity analysis, we performed a subgroup analysis of participants with both QFT and tuberculin skin test (TST) results. We defined “confirmed TB infection” when both TST and QFT results were positive and “no TB infection” if both TST and QFT results were negative. Participants with discordant TST and QFT results (i.e., TST negative and QFT positive, TST positive and QFT negative) were classified as “any discordance.”

Other important covariates, including age, sex, race, educational attainment, income to poverty ratio, country of birth, body mass index (BMI), diabetes mellitus status, HIV status, lipid profile, self-reported smoking behavior, alcohol consumption, statin prescription, and previous diagnosis of coronary heart disease, myocardial infarction, or stroke were also abstracted. We classified BMI as “underweight” (BMI <18.5 kg/m^2^), “normal” (BMI 18.5 – 24.9 kg/m^2^), “overweight” (BMI 25 – 29.9 kg/m^2^), and obese (BMI ≥30kg/m^2^). ^26^ As NHANES grouped individuals aged ≥80 years in one category, we divided age into quartile ranges and grouped as “quartile 1 (18 – 31 years)”, “quartile 2 (32 – 47 years)”, “quartile 3 (48 – 62 years)”, and “quartile 4 (≥63 years)” to account for the non-linearity of age in sensitivity analyses.

### Statistical Analysis

We estimated weighted prevalence and 95% confidence intervals (CI) to determine the burden of TBI and hypertension in the US adult population. Rao-Scott Chi-square tests were used to assess the bivariate association between participants’ demographic and clinical characteristics, TBI, Ag-NIL values, and hypertension. Multivariable robust Poisson regression with quasi-likelihood was used to estimate the association between TBI and hypertension, expressed in prevalence ratios (PRs) and 95% CI. The same regression approach was used to estimate the association between Ag-NIL responses and hypertension. In addition to prevalence ratios, we also estimated prevalence differences (PDs) and their 95%CI. Covariates included in the multivariable models were based on bivariate associations (Table S1 and S2), directed acyclic graphs ^27^, and established risk factors reported in previously published studies. We also assessed interaction between TBI and hypertension by participant characteristics (i.e., age, BMI, glycemic status, smoking status) on the additive (prevalence difference) and multiplicative (prevalence ratio) scales. All analyses were performed using *survey* package in R and accounted for the weighted stratified probability sample design of NHANES with a two-sided p-value less than 0.05 considered statistically significant.

### Subgroup and Sensitivity Analyses

Subgroup analyses were performed among those with previously diagnosed hypertension to determine the association between TBI (including Ag-NIL values) and controlled hypertension. Sensitivity analyses were performed to quantify systematic errors due to a) TBI misclassification, b) covariate misspecification in multivariable models, and c) the classification of age as a confounder. To address error resulting from TBI misclassification, we ran additional models with confirmed TB infection as the exposure. To quantify errors due to covariate misspecification, we ran multiple robust Poisson models with different sets of covariates and observed changes in prevalence ratios estimates across models. To account for the confounding effect of age, we ran multiple iterations of robust Poisson models with different forms of age measures (i.e., continuous and age quartiles).

## RESULTS

### Study population

In NHANES 2011 – 2012, 9,338 participants were surveyed and examined, 60.1% (5,615/9,338) of whom were ≥18 years old (Figure 1). Among included adults, 259 did not have valid blood pressure measurements. Of those with valid blood pressure measurements, 32 had a previous diagnosis of TB disease and 335 had a missing QFT, with 4,989 participants meeting eligiblity for this analytic cohort. The weighted prevalence of TBI in the cohort was 5.7% (95% confidence interval [CI] 4.7– 6.7) and any hypertension was present for 48.9% (95%CI 45.2 – 52.7) of participants (Table 1).

**Table 1.**
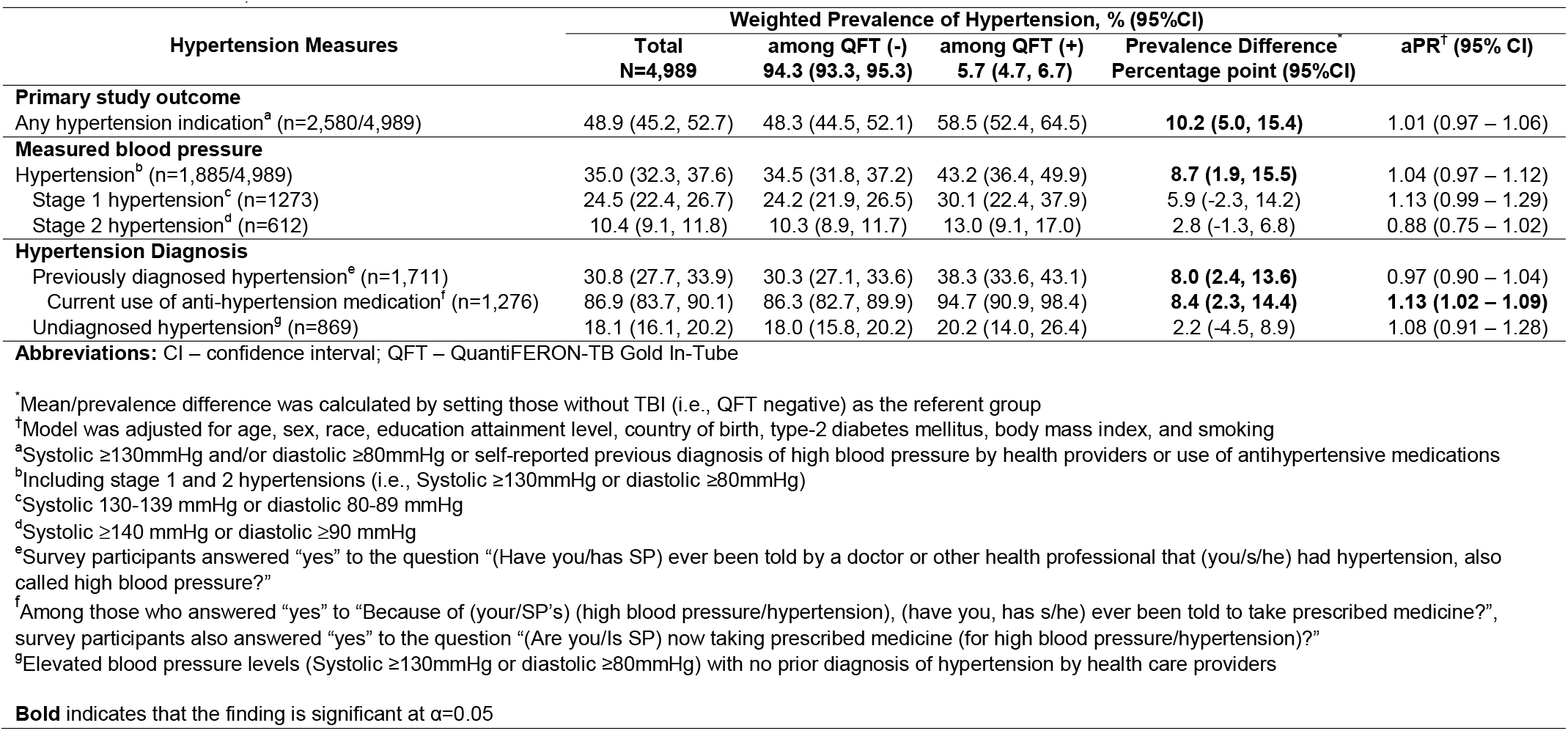
Weighted prevalence and adjusted prevalence ratios of hypertension measures by QuantiFERON-TB Gold In-Tube status among US adults, NHANES 2011-2012 *This table shows the prevalence of select hypertension measures in the overall adult cohort of NHANES 2011 – 2012 as well as stratified by their tuberculosis infection status. The crude measure of association was expressed as prevalence difference (PD), while the adjusted measure of association was expressed as prevalence ratio (PR)*.

| Hypertension Measures | Weighted Prevalence of Hypertension, % (95%CI) |  |  |  | aPR<sup>†</sup> (95% CI) |
| --- | --- | --- | --- | --- | --- |
|  | Total<br>N=4,989 | among QFT (-)<br>94.3 (93.3, 95.3) | among QFT (+)<br>5.7 (4.7, 6.7) | Prevalence Difference<sup>*</sup><br>Percentage point (95%CI) |  |
| **Primary study outcome** |  |  |  |  |  |
| Any hypertension indication<sup>a</sup> (n=2,580/4,989) | 48.9 (45.2, 52.7) | 48.3 (44.5, 52.1) | 58.5 (52.4, 64.5) | **10.2 (5.0, 15.4)** | 1.01 (0.97 – 1.06) |
| **Measured blood pressure** |  |  |  |  |  |
| Hypertension<sup>b</sup> (n=1,885/4,989) | 35.0 (32.3, 37.6) | 34.5 (31.8, 37.2) | 43.2 (36.4, 49.9) | **8.7 (1.9, 15.5)** | 1.04 (0.97 – 1.12) |
| Stage 1 hypertension<sup>c</sup> (n=1273) | 24.5 (22.4, 26.7) | 24.2 (21.9, 26.5) | 30.1 (22.4, 37.9) | 5.9 (-2.3, 14.2) | 1.13 (0.99 – 1.29) |
| Stage 2 hypertension<sup>d</sup> (n=612) | 10.4 (9.1, 11.8) | 10.3 (8.9, 11.7) | 13.0 (9.1, 17.0) | 2.8 (-1.3, 6.8) | 0.88 (0.75 – 1.02) |
| **Hypertension Diagnosis** |  |  |  |  |  |
| Previously diagnosed hypertension<sup>e</sup> (n=1,711) | 30.8 (27.7, 33.9) | 30.3 (27.1, 33.6) | 38.3 (33.6, 43.1) | **8.0 (2.4, 13.6)** | 0.97 (0.90 – 1.04) |
| Current use of anti-hypertension medication<sup>f</sup> (n=1,276) | 86.9 (83.7, 90.1) | 86.3 (82.7, 89.9) | 94.7 (90.9, 98.4) | **8.4 (2.3, 14.4)** | **1.13 (1.02 – 1.09)** |
| Undiagnosed hypertension<sup>g</sup> (n=869) | 18.1 (16.1, 20.2) | 18.0 (15.8, 20.2) | 20.2 (14.0, 26.4) | 2.2 (-4.5, 8.9) | 1.08 (0.91 – 1.28) |

**Abbreviations:** CI – confidence interval; QFT – QuantiFERON-TB Gold In-Tube
<sup>\*</sup>Mean/prevalence difference was calculated by setting those without TBI (i.e., QFT negative) as the referent group
<sup>†</sup>Model was adjusted for age, sex, race, education attainment level, country of birth, type-2 diabetes mellitus, body mass index, and smoking
<sup>a</sup>Systolic $\geq 130$ mmHg and/or diastolic $\geq 80$ mmHg or self-reported previous diagnosis of high blood pressure by health providers or use of antihypertensive medications
<sup>b</sup>Including stage 1 and 2 hypertensions (i.e., Systolic $\geq 130$ mmHg or diastolic $\geq 80$ mmHg)
<sup>c</sup>Systolic 130-139 mmHg or diastolic 80-89 mmHg
<sup>d</sup>Systolic $\geq 140$ mmHg or diastolic $\geq 90$ mmHg
<sup>e</sup>Survey participants answered “yes” to the question “(Have you/has SP) ever been told by a doctor or other health professional that (you/s/he) had hypertension, also called high blood pressure?”
<sup>f</sup>Among those who answered “yes” to “Because of (your/SP’s) (high blood pressure/hypertension), (have you, has s/he) ever been told to take prescribed medicine?”, survey participants also answered “yes” to the question “(Are you/Is SP) now taking prescribed medicine (for high blood pressure/hypertension)?”
<sup>g</sup>Elevated blood pressure levels (Systolic $\geq 130$ mmHg or diastolic $\geq 80$ mmHg) with no prior diagnosis of hypertension by health care providers
**Bold** indicates that the finding is significant at $\alpha=0.05$

### Associations between tuberculosis infection and hypertension

The prevalence of any hypertension was higher among those with TBI (58.5%, 95% CI 52.4 – 64.5) than those without TBI (48.3%, 95%CI 44.5 – 52.1) (prevalence difference [PD] 10.2%, 95%CI 5.0 – 15.4) (Table 1). After adjusting for potential confounders including age (continuous), sex, race, educational attainment level (as a proxy of socioeconomic status), country of birth, diabetes mellitus status, BMI, and smoking status, the prevalence of any hypertension was similar among those with and without TBI (adjusted prevalence ratio [aPR] 1.0, 95%CI 1.0 – 1.1). The association between TBI and hypertension was similar when examining the two components used to define our primary outcome (i.e., measured hypertension and self-reported hypertension/use of antihypertensive medications) both in the crude and adjusted models (Table 1).

### Association between Ag-NIL values and hypertension

The prevalence of any hypertension was highest among those with TBI and high Ag-NIL values (60.4%, 95%CI 53.0 – 67.7) compared to those with TBI and low Ag-NIL values (57.6%, 95%CI 48.7 – 66.6) or those without TBI (48.3%, 95%CI 44.5 – 52.1) (Table S3). After adjusting for age and gender, however, the prevalence of any hypertension was similar among the three QFT groups being compared (Table S4). Similar trends were also observed for the associations between Ag-NIL values and both measured hypertension and self-reported previous diagnosis of hypertension (Figure 2).

**Figure 2.**
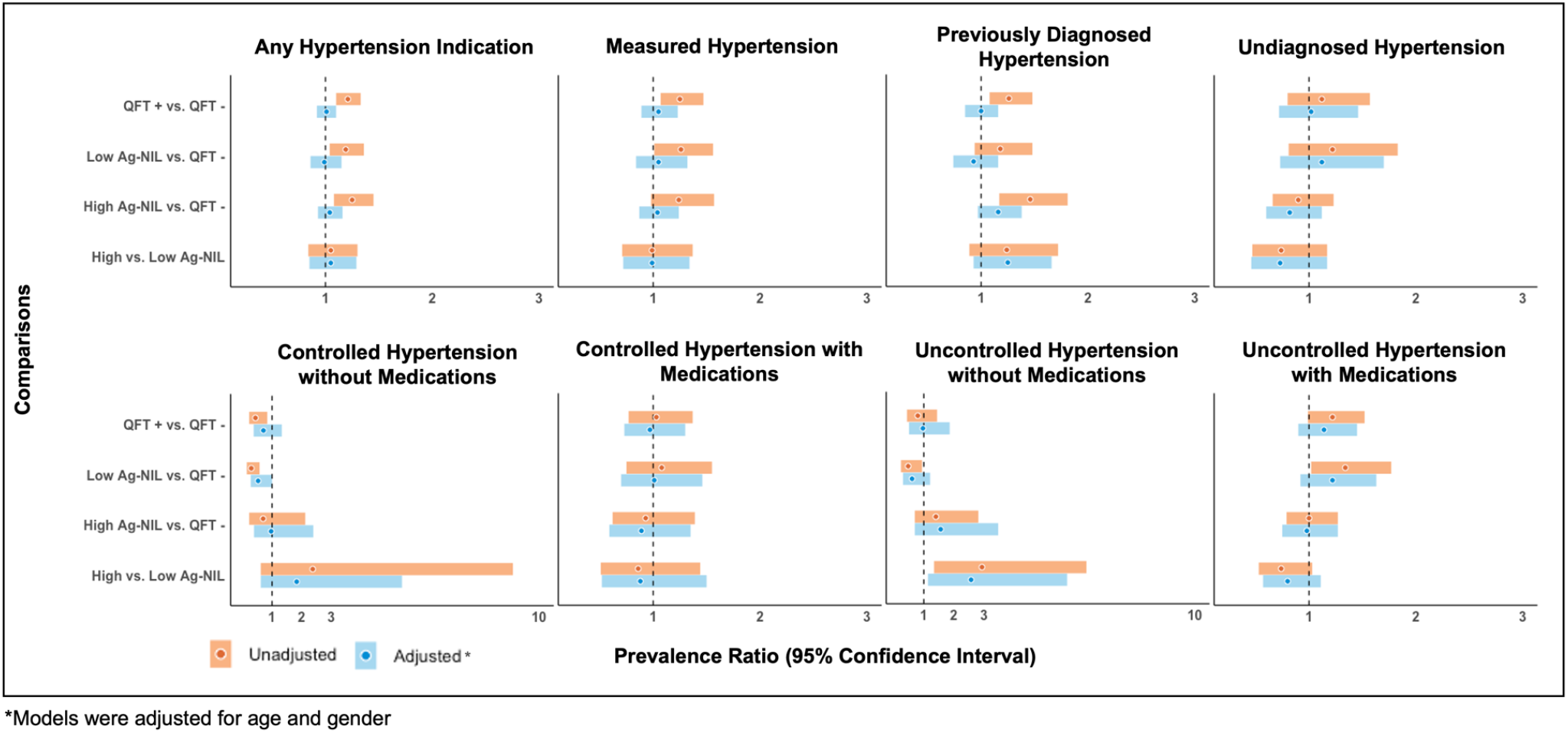
Crude and adjusted associations between QuantiFERON-TB Gold In-Tube results and select hypertension measures among US adults, NHANES 2011 – 2012 *Circles in this panel of figures indicate point estimates from the robust Poisson models, expressed as prevalence ratios with the colored bands indicating the accompanying 95% confidence intervals. The vertical dashed line on the x axis value of 1 marks the study null value (i*.*e*., *β estimates=0 or prevalence ratio=1*.*00), suggesting no association. The top panel figures were produced from analyses performed among eligible participants (n=4,989). The lower panel figures were produced from analyses performed among a subset of participants with known hypertension indication(n=1,711)*

### Interaction analyses: established hypertension risk factors and HIV

We observed relationships between TBI and hypertension among participants without established hypertension risk factors who would be considered at lower risk for CVD. For example, comparing individuals with with and without TBI, the prevalence of any hypertension was substantially higher among those with normal BMI (prevalence difference [PD] 17.7, 95%CI 6.3 – 29.2), euglycemia (PD 11.3, 95%CI 3.0 – 18.9), and non-smoking (PD 14.4, 95%CI 4.2 – 24.5) groups (Table 2). Product terms for BMI, glycemic level, and smoking status were non-significant on the prevalence ratio scale (p<0.05).

**Table 2.**
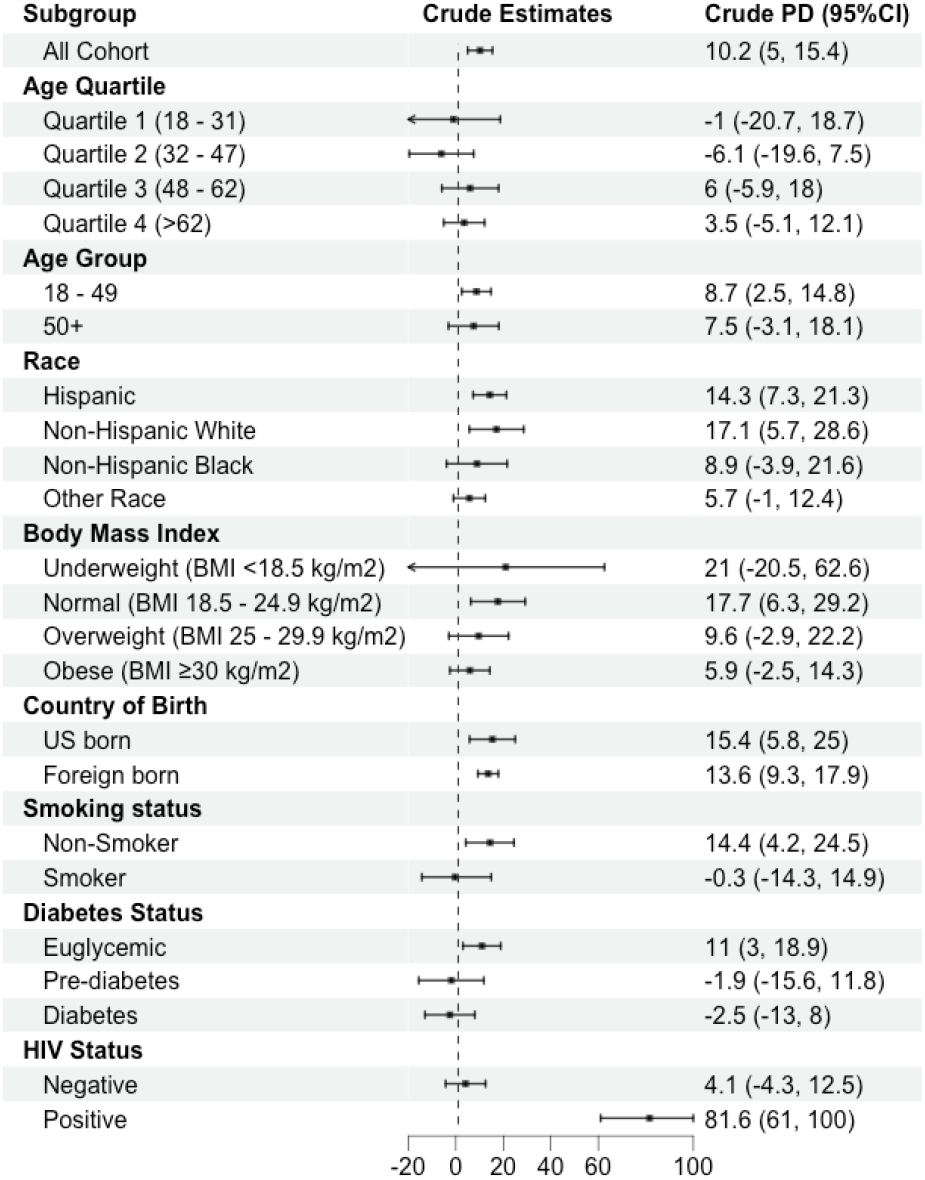
Relationship between positive QuantiFERON-TB result and hypertension: Stratified by demographic and clinical characteristics among US adults, NHANES 2011 – 2012 *This table shows results from the analyses with statistical interaction term included in the robust Poisson models to evaluate the joint effect between tuberculosis infection and other key risk factors on hypertension. We selected these “moderator” variables by identifying common risk factors for cardiovascular diseases from published studies (e*.*g*., *age, race, body mass index, country of birth, smoking status, diabetes status, and HIV status*.

We also found that the association between TBI and hypertension was significantly different across HIV status. For instance, the prevalence difference of any hypertension comparing those with TBI to those without TBI was 4.1 percentage points (95%CI -4.3 – 12.5) among those without HIV infection and 81.6 percentage points (95%CI 61.0 – 100.0) among those with HIV infection. After adjusting for age and gender, the adjusted prevalence ratio was 0.9 (95%CI 0.8 – 1.1) among those without HIV infection and 6.2 (95%CI 1.8 – 21.7) among those with HIV infection (statistical interaction p<0.01) (Table S5).

### Subgroup and sensitivity analyses

From subgroup analyses conducted among those with known hypertension, the prevalence of controlled hypertension without medications was significantly lower among those with positive QFT (5.2%, 95%CI 2.0 – 8.3) compared to those with negative QFT (11.8%, 95%CI 9.5 – 14.0), although the association was no longer significant after adjusting for key confounders (aPR 0.6, 95%CI 0.4 – 1.1) (Table 3). Conversely, the prevalence of uncontrolled hypertension with medications, the more severe form of hypertension, although non-significant, were slightly higher among those with positive QFT compared to those with negative QFT (Figure 2).

**Table 3.**
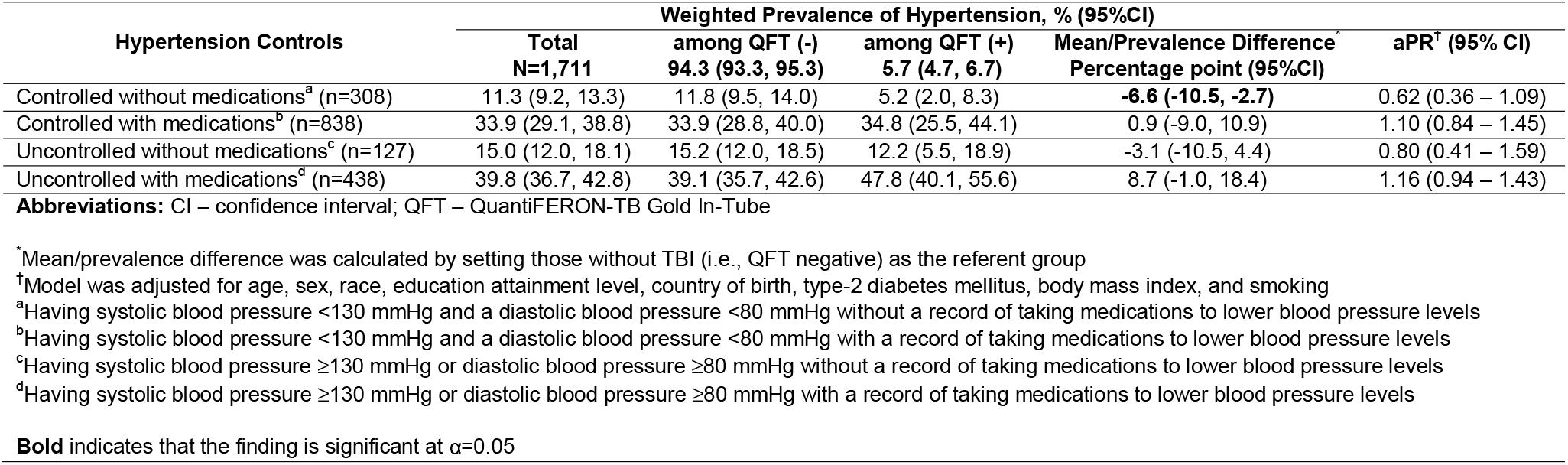
Weighted prevalence and adjusted prevalence ratios of controlled and uncontrolled hypertension by QuantiFERON-TB Gold In-Tube status among US adults with known hypertension, NHANES 2011-2012 *This table summarizes findings on whether latent tuberculosis infection is associated with severe clinical manifestation of hypertension, indicated by elevated measured blood pressure levels with the use of antihypertensive medications among individuals with known hypertension indications (n=1,711)*

In models with confirmed TB infection (i.e., positive QFT and positive TST) as the study exposure, the prevalence of any hypertension was highest among those with confirmed TB infection (60.8%, 95%CI 51.4 – 70.3) compared to those with no TB infection (49.6%, 95%CI 45.7 – 53.5) or those with discordant TST and QFT results (52.7%, 95%CI 43.9 – 61.6) (p=0.12) (Table S6). We observed similar trends in the crude and adjusted associations between TBI and hypertension when we used both QFT and TST (Table S7) vs. QFT alone to define TBI. Results from sensitivity analyses to quantify bias due to covariate misspecification in the multivariable models indicated that prevalence ratios of any hypertension comparing those with positive QFT to those with negative QFT were similar when age was treated continuously or grouped in quartiles (ranged from 1.0 – 1.1) (Table S8).

## DISCUSSION

Using data representative of US adult population, we found a high prevalence of hypertension (i.e., nearly 1 out of 2) in the 2011 – 2012 NHANES cycle. We reported similar adjusted prevalence of hypertension among individuals with or without TBI. In our study, individuals with positive QFT and high Ag-NIL values were more likely to have any hypertension, but less likely to have the more severe form of hypertension (i.e., uncontrolled hypertension without medications). We also observed that the association between TBI and hypertension was more common among individuals without established hypertension risk factors. Collectively, our results provide preliminary epidemiologic evidence suggesting that hypertension, a well-established intermediate for CVD, was more common among individuals with TBI than those without TBI in the US populations.

Our finding suggesting that hypertension is more common among individuals with TBI than those without TBI is consistent with previous studies. For example, a retrospective cohort study conducted among 5,185 individuals with TBI and healthy controls using data from a large metropolitan healthcare system in the US reported a higher hazard rates of hypertension incidence (defined by ICD-9 codes) among those with TBI (defined by ICD-9 codes and tuberculin skin test/IFN-γ release assay) compared to healthy controls without TBI (HR 2.0, 95%CI 1.6 – 2.5). ^5^ In addition, a cross-sectional study conducted among 2,351 TST-positive individuals in South India reported a slightly higher prevalence of hypertension (defined as systolic >130 mmHg) among those with confirmed TBI (defined as TST and QFT positive) (15%) compared to those latent TB negative (12%) (aOR 1.18, 95%CI 1.0 – 1.56). ^28^ Unlike the two studies mentioned above, we used a more comprehensive definition of hypertension by combining objectively measured blood pressure levels (systolic and diastolic) and known hypertension indications (i.e., previous hypertension diagnosis or self-reported use of antihypertensive medications) to avoid potential misclassification.

Furthermore, we also reported that the prevalence of hypertension was highest among individuals with positive QFT and high Ag-NIL values, but we observed no dose-response relationship nor statistical significance after adjusting for key risk factors. TB infection has been associated with enhanced levels of systemic inflammation and immune activation, including increased expression of tumor necrosis factor (TNF)-α, interferons, and interlukin-6 (IL-6) ^4-7^.

These chemokines and dysfunctional immune responses play an important role in the pathogenesis of hypertension and CVD ^29 30^. Individuals with positive QFT and higher Ag-NIL values are more likely to develop to active TB ^23 31^ as they may have higher mycobacterial burden, ^21^ and thus, could potentially have higher degree of inflammation or immune responses to the bacterial infection.

Our cross-sectional study design may not be the appropriate design to observe the expected associations or dose-response relationship between TBI, IFN-γ TB antigen responses, and hypertension. Furthermore, the time of TBI in the life-course may have different implications on TBI and hypertension association. In this NHANES cohort, the majority (>90%) of foreign born with positive QFT have stayed in the US for ≥5 years, and thus, we postulated that TBI happened before arriving in the US. It is plausible that these individuals are either in the latent or incipient stage where there is no to minimum bacteria replication, and thus, minimum pro-inflammatory responses. ^32^ Prospective studies to follow individuals with recent TBI diagnosis are still warranted to determine the hypertension and CVD risk trajectories.

Interestingly, we observed associations between TBI and hypertension among those with normal BMI, euglycemic, and non-smokers. These groups may be considered at lower risk of CVD. This finding further reinforces the premise that there is likely to be differing effects of TBI on hypertension risk within subgroups. Further investigations and modeling studies are needed to determine whether targeted TB preventive treatment is effective to reduce the global burden of CVD among these groups.

Last, we reported that HIV infection may modify the association between TBI and hypertension. However, this finding needs to be interpreted with caution considering the low prevalence of HIV infection in the 2011-2012 NHANES cycle. Previous studies demonstrated that hypertension is more common among individuals with HIV infection on antiretroviral therapy compared to those without HIV infection, ^33 34^ and that there are several plausible pathways regarding how HIV infection could lead to hypertension. ^33^ For example, the chronic inflammation among people living with HIV (PLWH), even among those with undetectable viral loads on stable antiretroviral therapy, would trigger host immune activation (e.g., upregulation of IL-6) and could lead to stiff blood vessels and impact hypertension risk. ^35 36^ Further clinical studies are warranted to fully assess the joint effect between HIV (including HIV clinical characteristics) and TBI, and its association with hypertension.

Our study is subject to limitations. First, our TBI definition (i.e., according to QFT positivity) may include a broad spectrum of individuals who may have cleared the infection, have latent TB, incipient TB, or even subclinical TB since no further clinical assessment was made (e.g., chest X-ray). ^37^ Second, we could not determine the temporal relationship between TBI and hypertension with the cross-sectional study design used in the present paper. Third, hypertension is known to be multifactorial, and we did not account for other key variables that could potentially affect blood pressure level including stress, family history, diet (e.g., sodium intake), lifestyle (e.g., physical activity), geographical delineation (i.e., rural vs. urban), or illicit drug use. If some of these variables are associated with TBI, it is plausible that our reported estimates are slightly distorted due to residual confounding effects. Additionally, we did not account for any record of hypertension prescription, or other commonly prescribed medications that could potentially affect blood pressure levels. Fourth, we defined some of our key variables (including hypertension status and hypertension medication intake) with self-reported information that may be prone to recall bias and likely included some misclassification.

However, if misclassification of hypertension was non-differential with respect to TBI, we expect any misclassification in our results would likely biased towards the null ^38^. Fourth, we did not take into consideration the CD4 count for the HIV-stratified analyses due to the small, unweighted frequency of individuals with HIV infection. Last, this study was conducted using survey data representative of US adult population but may not be generalizable to other regions with higher TB burdens.

In conclusion, we reported a higher prevalence of hypertension among individuals with positive QFT, although the association was non-significant after adjusting for key confounders, particularly age. To determine the direction of the association between TBI and hypertension, a prospective study following hypertension-free individuals at TBI diagnosis is warranted and would help establish the biological pathways regarding how TBI might increase the risk of CVD. Importantly, our results underscore the need to screen for hypertension and other metabolic disorders among those with TBI, especially among those without traditional CVD risk factors; doing so may help prevent premature deaths attributed to TB and CVD.

## Data Availability

Data used for this analysis is publicly available for download in US CDC National Health and Nutrition Examination Survey's website.

https://www.cdc.gov/nchs/nhanes/index.htm

## DECLARATIONS AND ACKNOWLEDGMENTS

### Author contributions

MAH, MJM, and ADS conceived the study design. ADS performed the analyses. ADS, MAH, and MJM wrote the first draft of the manuscript. SCA, UPG, EMU, and JRA assisted with further drafting and revisions of manuscripts. All authors reviewed and approved the final version of the manuscript.

