## Supplemental Materials for "Tuberculosis infection and hypertension: Prevalence estimates from the US National Health and Nutrition Examination Survey"

**SUPPEMENTAL MATERIALS**

|  |  | **Page(s)** |
| --- | --- | --- |
| **Table S1** | Weighted prevalence of and characteristics associated with tuberculosis infection among according to QuantiFERON-TB Gold In-Tube results among representative of civilian, non-institutionalized US adult population, NHANES 2011-2012 | i – iv |
| **Table S2** | Weighted prevalence of and characteristics associated with hypertension among representative of civilian, non-institutionalized US adult population, NHANES 2011-2012 | v – viii |
| **Table S3** | Weighted prevalence of various hypertension classifications by interferon gamma tuberculosis antigen responses among representative of civilian, non-institutionalized US adult population, NHANES 2011-2012 | ix – x |
| **Table S4** | Crude and adjusted associations between interferon gamma tuberculosis antigen responses and hypertension among representative of civilian, non-institutionalized US adult population, NHANES 2011 – 2012 | xi – xii |
| **Table S5** | The crude and adjusted prevalence odds ratios of any hypertension stratified by race, body mass index category, and foreign-born status, among representative of civilian, non-institutionalized US adult population, NHANES 2011 – 2012 | xiii – xiv |
| **Table S6** | Weighted prevalence of various hypertension classifications by confirmed tuberculosis infection status among representative of civilian, non-institutionalized US adult population, NHANES 2011-2012 | xv – xvi |
| **Table S7** | Crude and adjusted associations between confirmed tuberculosis infection status and hypertension among representative of civilian, non-institutionalized US adult population, NHANES 2011 – 2012 | xvii – xviii |
| **Table S8** | Sensitivity analysis to account for misclassification of covariates and different ways to handle age (confounder) included in the multivariable survey-weighted robus Poisson models to estimate the association between tuberculosis infection and hypertension among representative of civilian, non-institutionalized US adult population, NHANES 2011-2012 | xix – xx |

**Supplemental Materials**

**Table S1.** Weighted prevalence of and characteristics associated with tuberculosis infection among according to QuantiFERON-TB Gold In-Tube results among representative of civilian, non-institutionalized US adult population, NHANES 2011-2012

| **Characteristics** | **Weighted Prevalence, % (95%CI)** | | | **P-Values**  **(X^2^)^†^** |
| --- | --- | --- | --- | --- |
|  | **QFT Negative**  **% (95% CI)**  94.3 (93.3 – 95.3) | **QFT Positive**  **% (95% CI)**  5.7 (4.7 – 6.7) | **Mean/Prevalence Difference^*^**  **Percentage point (95%CI)** |  |
| Any hypertension indication^a^  No  Yes | 95.4 (94.4 – 96.4)  93.2 (91.8 – 94.5) | 4.6 (3.6 – 5.6)  6.8 (5.5 – 8.2) | Reference  2.2 (1.0, 3.4) | **<0.001** |
| Age, years  Mean (95%CI)  Age groups  Quartile 1 (18 – 31)  Quartile 2 (32 – 47)  Quartile 3 (48 – 62)  Quartile 4 (>62)  18 – 49  ≥50 | 46.0 (44.1 – 48.0)  97.2 (96.2 – 98.3)  95.5 (94.4 – 96.6)  92.0 (89.2 – 94.7)  91.9 (89.8 – 94.1)  94.9 (94.1 – 95.7)  92.5 (90.4 – 94.7) | 53.2 (51.2 – 55.1)  2.8 (1.7 – 3.8)  4.5 (3.4 – 5.6)  8.0 (5.3 – 10.8)  8.1 (5.9 – 10.2)  5.1 (4.3 – 5.9)  7.5 (5.3 – 9.6) | 7.1 (5.1, 9.2)  Reference  1.7 (0.1, 3.3)  5.3 (2.1, 8.4)  5.3 (3.4, 7.2)  Reference  2.4 (0.5, 4.2) | **<0.001^‡^**  **<0.001**  **0.004** |
| Sex  Male  Female | 93.4 (92.1 – 94.6)  95.2 (94.1 – 96.2) | 6.6 (5.4 – 7.9)  4.8 (3.8 – 5.9) | Reference  -1.8 (-3.2, -0.4) | **0.005** |
| Race  Hispanic  Non-Hispanic white  Non-Hispanic black  Other race | 87.6 (85.4 – 89.9)  96.8 (95.8 – 97.8)  92.8 (90.9 – 94.7)  86.7 (84.0 – 89.5) | 12.4 (10.1 – 14.6)  3.2 (2.2 – 4.2)  7.2 (5.3 – 9.1)  13.3 (10.5 – 16.0) | Reference  -9.2 (-12.0, -6.4)  -5.1 (-7.7, -2.6)  0.9 (-2.4, 4.2) | **<0.001** |
| Education (n=4,757)  Less than 9^th^ grade  9-11^th^ grade  High school graduate  Some college  College graduate or above    *Missing (n=264)* | 82.4 (77.8 – 86.9)  92.4 (90.1 – 94.7)  92.9 (90.5 – 95.3)  96.7 (95.4 – 98.0)  95.0 (93.4 – 96.7)  98.0 (95.6 – 100.0) | 17.6 (13.1 – 22.2)  7.6 (5.3 – 9.9)  7.1 (4.7 – 9.5)  3.3 (2.0 – 4.6)  5.0 (3.2 – 6.6)  2.0 (0 – 4.4) | Reference  -10.4 (-15.5, -4.6)  -10.6 (-15.8, -5.3)  -14.3 (-19.2, -9.5)  -12.7 (-17.1, -8.3) | **<0.001** |
| Ratio of family income to poverty (n=4,623)  Mean (95%CI)  0 – 0.99  1 – 1.99  2 – 2.99  3 – 3.99  4 – 4.99  ≥5  *Missing (n=396)* | 2.9 (2.7 – 3.1)  92.0 (89.8 – 94.2)  92.5 (91.0 – 94.1)  94.9 (91.7 – 98.1)  95.8 (94.0 – 97.6)  96.7 (94.5 – 98.8)  95.9 (94.1 – 97.7)  91.9 (88.4 – 95.5) | 2.4 (2.1 – 2.7)  8.0 (5.8 -10.2)  7.5 (5.9 – 9.0)  5.1 (1.9 – 8.3)  4.2 (2.4 – 6.0)  3.3 (1.2 – 5.5)  4.1 (2.3 – 5.9)  8.1 (4.5 – 11.6) | -0.5 (-0.9, -0.2)  Reference  -0.5 (-3.1, 2.1)  -2.9 (-7.0, 1.2)  -3.8 (-6.4, -1.3)  -4.7 (-8.3, -1.1)  -3.9 (-6.9, -0.9) | **0.005^‡^**  **0.004** |
| Foreign born (n=4,987)  No  Yes  *Missing (n=2)* | 96.5 (95.5 – 97.6)  83.6 (80.8 – 86.3)  100.0 (100.0 – 100.0) | 3.5 (2.4 – 4.5)  16.4 (14.0 – 19.2)  0 (0 – 0) | Reference  13.0 (9.6, 16.3) | **<0.001** |
| BMI, kg/m^2^ (n=4,930)  Mean (95%CI)    BMI categories  Underweight (<18.5 kg/m^2^)  Normal (18.5 – 24.9 kg/m^2^)  Overweight (25 – 29.9 kg/m^2^)  Obese (≥30 kg/m^2^)  *Missing (n=59)* | 28.7 (28.2 – 29.1)  93.1 (87.8 – 98.4)  93.9 (92.0 – 95.8)  94.5 (93.2 – 95.9)  94.4 (93.2 – 95.5)  95.8 (90.2 – 100.0) | 28.9 (27.8 – 30.1)  6.9 (1.6 – 12.2) 6.1 (4.2 – 8.0)  5.5 (4.1 – 6.8)  5.6 (4.5 – 6.8)  4.2 (0 – 9.8) | 0.2 (-0.7, 1.2)  0.9 (-3.7, 5.4)  Reference  -0.6 (-2.6, 1.4)  -0.4 (-2.6, 1.7) | 0.603^‡^  0.868 |
| Smoking status (n=4,722)  Never smokers^b^  Past smokers^c^  Current smokers^d^  *Missing (n=267)* | 94.9 (94.0 – 95.8)  93.1 (90.7 – 95.4)  93.3 (90.7 – 95.8)  98.1 (95.7 – 100.0) | 5.1 (4.2 – 6.0)  6.9 (4.6 – 9.3)  6.7 (4.2 – 9.3)  1.9 (0 – 4.3) | Reference  1.8 (-0.7, 4.3)  1.6 (-1.0, 4.2) | 0.145 |
| Heavy alcohol drinking (n=3,867)  No  Yes^e^  *Missing (n=1,122)* | 95.0 (93.5 – 96.4)  94.7 (93.7 – 95.8)  92.0 (90.5 – 93.6) | 5.0 (3.6 – 6.5)  5.3 (4.2 – 6.3)  8.0 (6.4 – 9.5) | Reference  0.3 (-1.1, 1.6) | 0.686 |
| HbA1c, %  Mean (95%CI)  Diabetes categories^f^  Normal  Prediabetes  Diabetes | 5.6 (5.6 – 5.7)  95.5 (94.6 – 96.4)  93.4 (91.7 – 95.0)  88.9 (85.2 – 92.5) | 5.9 (5.7 – 6.0)  4.5 (3.6 – 5.4)  6.6 (5.0 – 8.3)  11.1 (7.5 – 14.8) | 0.3 (0.1, 0.4)  Reference  2.1 (0.8, 3.5)  6.6 (2.9, 10.4) | **0.001^‡^**  **<0.001** |
| HIV co-infection status (n=3,408)  Negative  Positive  *Missing (n=1,600)* | 95.4 (94.4 – 96.4)  96.1 (88.3 – 100.0)  91.3 (89.3 – 93.3) | 4.6 (3.6 – 5.6)  3.9 (0 – 11.7)  8.7 (6.7 – 10.7) | Reference  0.7 (-7.0, 8.3) | 0.861 |
| ***Dyslipidemia Measures*** |  |  |  |  |
| HDL (mg/dL) (n=4,889)  Mean (95%CI)  HDL levels^g^  Normal  Lower  *Missing (n=100)* | 52.8 (51.8 – 53.9)  94.6 (93.5 – 95.7)  93.6 (92.4 – 94.9)  91.8 (82.9 – 100.0) | 51.7 (48.9 – 54.5)  5.4 (4.3 – 6.5)  6.4 (5.1 – 7.6)  8.2 (0 – 17.1) | -1.1 (-3.5, 1.2)  Reference  1.0 (-0.3, 2.2) | 0.330^‡^  0.113 |
| LDL^h^ (mg/dL) (n=2,236)  Mean (95%CI)  LDL levels  Normal (<130 mg/dL)  Elevated (130 – 159 mg/dL)  High (≥160 mg/dL)  *Missing (n=67)* | 114.8 (112.5 – 117.0)  94.3 (92.8 – 95.8)  95.8 (94.6 – 97.2)  94.5 (90.7 – 98.4)  99.5 (98.3 – 100.0) | 113.1 (107.1 – 119.2)  5.7 (4.2 – 7.2)  4.2 (2.8 – 5.6)  5.4 (1.6 – 9.3)  0.5 (0 – 1.7) | -1.6 (-8.4, 5.1)  Reference  -1.5 (-3.3, 0.4)  -0.2 (-3.9, 3.4) | 0.614^‡^  0.397 |
| Total Cholesterol (mg/dL) (n=4,889)  Mean (95%CI)  Total cholesterol levels  Low (≤130 mg/dL)  Normal (131 – 199 mg/dL)  Elevated (≥200 mg/dL)  *Missing (n=100)* | 194.2 (191.9 – 196.4)  93.3 (89.8 – 96.8)  94.5 (93.3 – 95.7)  94.2 (82.9 – 100.0)  91.8 (82.9 – 100.0) | 196.8 (192.5 – 201.0)  6.7 (3.2 – 10.2)  5.5 (4.2 – 6.7)  5.8 (4.4 – 7.2)  8.2 (0 – 17.1) | 2.6 (-1.3, 6.5)  Reference  -1.3 (-5.6, 3.0)  -0.9 (-4.9, 3.1) | 0.183^‡^  0.728 |
| Triglyceride^h^ (mg/dL) (n=2,276)  Mean (95%CI)  Triglyceride levels  Optimal (<150 mg/dL)  Elevated (150 – 199 mg/dL)  High (≥200 mg/dL)  *Missing (n=27)* | 129.6 (118.9 – 140.2)  94.6 (93.0 – 96.2)  94.9 (92.5 – 97.2)  95.4 (93.6 – 97.2)  100.00 (100.0 – 100.0) | 123.4 (111.8 – 135.0)  5.4 (3.8 -7.0)  5.1 (2.8 – 7.5)  4.6 (2.8 – 6.4)  0 (0 – 0) | -6.2 (-20.5, 8.1)  Reference  -0.3 (-3.1, 2.6)  -0.9 (-3.2, 1.5) | 0.374^‡^  0.796 |
| Any dyslipidemia^i&h^ (n=2,277)  No  Yes  *Missing (n=26)* | 94.4 (92.1 – 96.7)  94.9 (93.6 – 96.2)  100.0 (100.0 – 100.0) | 5.6 (3.3 – 7.9)  5.1 (3.8 – 6.4)  0 (0 – 0) | Reference  -0.5 (-3.0, 2.0) | 0.637 |
| Statin prescription^j^ (n=2,770)  No  Yes  *Missing (n=2,238)* | 94.2 (92.7 – 95.6)  93.5 (91.8 – 95.2)  94.7 (93.6 – 95.8) | 5.8 (4.4 – 7.3)  6.5 (4.7 – 8.2)  5.3 (4.2 – 6.4) | Reference  0.6 (-1.3, 2.6) | 0.497 |
| CHD^k^ (n=4,712)  No  Yes  *Missing (n=277)* | 94.1 (93.0 – 95.1)  96.5 (94.7 – 98.3)  97.8 (95.5 – 100.0) | 5.9 (4.9 – 7.0)  3.5 (1.7 – 5.3)  2.2 (0 – 4.5) | Reference  -2.4 (-4.6, - 0.2) | **0.047** |
| Heart attack^l^ (n=4,723)  No  Yes  *Missing (n=266)* | 94.1 (93.1 – 95.1)  96.3 (94.5 – 98.1)  98.1 (95.7 – 100.0) | 5.9 (4.9 – 6.9)  3.7 (1.9 – 5.5)  1.9 (0 – 4.3) | Reference  -2.2 (-3.6, -0.8) | **0.008** |
| Stroke^m^ (n=4,725)  No  Yes  *Missing (n=264)* | 94.3 (93.2 – 95.3)  90.7 (86.4 – 94.9)  98.1 (95.7 – 100.0) | 5.7 (4.7 – 6.8)  9.3 (5.1 – 13.6)  1.9 (0 – 4.3) | Reference  3.6 (-0.9, 8.0) | **0.042** |
| Abbreviations:  BMI – body mass index; CHD – coronary heart disease; CI – confidence interval; HbA1c – glycated hemoglobin; HDL – high-density lipoprotein; HIV – human immunodeficiency virus; LDL – low-density lipoprotein; NHANES – National Health and Nutrition Examination Survey; QFT - QuantiFERON Gold-In-Tube;  ^*^Mean/prevalence difference was calculated by setting those without TBI (i.e., QFT negative) as the referent group, unless indicated otherwise (with “reference” statement)  ^†^P-values from Rao-Scott Chi-square tests, unless indicated otherwise  ^‡^P-values from t-tests  ^a^Systolic ≥130mmHg and/or diastolic ≥80mmHg or any previous diagnosis of high blood pressure by health providers  ^b^Survey participants answered “No” to the question “(Have you/has SP) smoked at least 100 cigarettes in life?  ^c^Survey participants answered “Not at all” to the question “(Do you/does SP) now smoke cigarettes?” and “Yes” to the question “(Have you/has SP) smoked at least 100 cigarettes in life?  ^d^Survey participants answered “Every day” or “Some days” to the question “(Do you/does SP) now smoke cigarettes?” and “Yes” to the question “(Have you/has SP) smoked at least 100 cigarettes in life?  ^e^Survey participants answered “Yes” to the question “Was there ever time or times in (your/SP’s) life when (you/he/she) drank 4 (for female) or 5 (for male) or more drinks of any kind of alcoholic beverage almost every day?”  ^f^Diabetes was categorized according to HbA1c levels and self-reported previous type-2 diabetes mellitus diagnosis by health care providers  ^g^HDL level was using gender-specific cut-offs: “normal” HDL was defined if HDL level was ≥40 mg/dL for male or ≥50 mg/dL for female; and “lower” HDL was defined if HDL level was <40 mg/dL for male or <50 mg/dL for female  ^h^LDL and triglyceride measurements were done among a subset of survey participants who were fasting and appropriate weight variable (for those who were fasting) was applied accordingly  ^i^Any dyslipidemia was defined as having either elevated LDL, total cholesterol, triglyceride, or lower HDL levels  ^j^Taken statin in the past 30 days prior to survey date, survey participants were also asked to show medicine container to surveyor/enumerator  ^k^Survey participants answered “Yes” to the question “Has a doctor or other health professional ever told (you/SP) that (you/s/he) had coronary heart disease?”  ^l^Survey participants answered “Yes” to the question “Has a doctor or other health professional ever told (you/SP) that (you/s/he) had a heart attack (also called myocardial infarction)?”  ^m^Survey participants answered “Yes” to the question “Has a doctor or other health professional ever told (you/SP) that (you/s/he) had a stroke?”  **Bold** indicates that the finding is statistically significant at α=0.05 | | | | |

**Table S2.** Weighted prevalence of and characteristics associated with hypertension among representative of civilian, non-institutionalized US adult population, NHANES 2011-2012

| **Characteristics** | **Weighted Prevalence, % (95%CI)** | | | **P-Values**  **(X^2^)^†^** |
| --- | --- | --- | --- | --- |
|  | **No Hypertension**  **% (95% CI)**  51.1 (47.4 – 54.8) | **Any Hypertension^a^**  **% (95% CI)**  48.9% (45.2 – 52.6) | **Mean/Prevalence Difference^*^**  **Percentage point (95%CI)** |  |
| QFT result  Negative  Positive | 51.7 (47.9 – 55.5)  41.5 (35.5 – 47.6) | 48.3 (44.5 – 52.1)  58.5 (52.4 – 64.5) | Reference  10.2 (5.0, 15.4) | **<0.001** |
| Age, years  Mean (95%CI)  Age group  Quartile 1 (18 – 31)  Quartile 2 (32 – 47)  Quartile 3 (48 – 62)  Quartile 4 (>62)  18 – 49  ≥50 | 38.9 (37.3 – 40.6)  80.8 (78.6 – 83.1)  57.5 (52.3 – 62.7)  38.0 (34.4 – 41.7)  23.0 (18.9 – 27.1)  57.1 (53.1 – 61.1)  34.2 (30.2 – 38.3) | 54.3 (52.8 – 55.7)  19.2 (16.9 – 21.4)  42.5 (37.3 – 47.7)  62.0 (58.3 – 65.6)  77.0 (72.9 – 81.1)  42.9 (38.9 – 46.9)  65.8 (61.7 – 69.8) | 15.3 (14.0, 16.6)  Reference  23.4 (18.6, 28.1)  42.8 (37.9, 47.7)  57.8 (53.1, 62.5)  Reference  22.9 (17.6, 28.2) | **<0.001**  **<0.001**  **<0.001** |
| Sex  Male  Female | 47.7 (43.2 – 52.2)  54.4 (50.4 – 58.4) | 52.3 (47.8 – 56.8)  45.6 (41.6 – 49.6) | Reference  -6.7 (-10.9, -2.5) | **0.001** |
| Race  Hispanic  Non-Hispanic white  Non-Hispanic black  Other race | 61.3 (55.8 – 66.8)  49.6 (44.7 – 54.4)  43.6 (39.9 – 47.4)  56.5 (51.5 – 61.5) | 38.7 (33.2 – 44.2)  50.4 (45.6 – 55.3)  56.4 (52.6 – 60.1)  43.5 (38.5 – 48.5) | Reference  11.7 (5.3, 18.2)  17.7 (11.8, 23.5)  4.8 (-2.2, 11.7) | **<0.001** |
| Education (n=4,725)  Less than 9^th^ grade  9-11^th^ grade  High school graduate  Some college  College graduate or above    *Missing (n=264)* | 39.0 (31.3 – 46.9)  42.3 (36.9 – 47.6)  45.5 (40.9 – 50.1)  51.7 (46.3 – 57.0)  55.3 (48.9 – 61.5)  86.7 (81.8 – 91.5) | 61.0 (53.3 – 68.7)  57.7 (52.3 – 63.1)  54.5 (49.9 – 59.1)  48.3 (42.9 – 53.7)  44.7 (38.4 – 51.1)  13.3 (8.5 – 18.2) | Reference  -3.2 (-13.4, 6.9)  -6.5 (-14.9, 2.0)  -12.7 (-20.9, -4.5)  -16.2 (-25.5, -7.0) | **<0.001** |
| Ratio of family income to poverty (n=4,593)  Mean (95%CI)  0 – 0.99  1 – 1.99  2 – 2.99  3 – 3.99  4 – 4.99  ≥5  *Missing (n=396)* | 2.8 (2.6 – 3.1)  55.8 (49.1 – 62.5)  49.6 (43.3 – 55.9)  49.4 (43.7 – 55.0)  53.5 (48.6 – 58.4)  47.6 (39.8 – 55.0)  50.9 (43.0 – 58.7)  49.4 (39.9 – 58.8) | 2.9 (2.7 – 3.1)  44.2 (37.5 – 50.9)  50.4 (44.1 – 56.7)  50.6 (45.0 – 56.3)  46.5 (41.6 – 51.4)  52.4 (44.6 – 60.2)  49.1 (41.3 – 57.0)  50.6 (41.2 – 60.1) | 0.1 (-0.1, 0.3)   Reference  6.2 (-0.5, 12.9)  6.4 (-2.4, 15.3)  2.3 (-4.9, 9.5)  8.2 (-2.7, 19.0)  4.9 (-3.5, 13.4) | 0.439  0.436 |
| Foreign born (n=5,019)  No  Yes  *Missing (n=2)* | 49.2 (45.9 – 52.6)  60.1 (54.7 – 65.5)  70.6 (8.7 – 100.0) | 50.8 (47.4 – 54.1)  39.9 (34.5 – 45.3)  29.4 (0 – 91.3) | Reference  -10.8 (-14.5, -7.2) | **<0.001** |
| BMI, kg/m^2^ (n=4,930)  Mean (95%CI)    BMI categories  Underweight (<18.5 kg/m^2^)  Normal (18.5 – 24.9 kg/m^2^)  Overweight (25 – 29.9 kg/m^2^)  Obese (≥30 kg/m^2^)  *Missing (n=59)* | 27.2 (26.7 – 27.8)  68.6 (61.2 – 76.0)  67.4 (62.8 – 72.0)  49.8 (46.2 – 53.4)  38.0 (33.6 – 42.5)  26.1 (7.3 – 45.0) | 30.2 (29.7 – 30.8)  31.4 (24.0 – 38.8)  32.6 (28.0 – 37.2)  50.2 (46.6 – 53.8)  62.0 (57.5 – 66.4)  73.9 (55.0 – 92.7) | 3.0 (2.4, 3.7)  -1.2 (-9.3, 6.9)  Reference  17.6 (14.4, 20.9)  29.4 (23.3, 35.5) | **<0.001**  **<0.001** |
| Smoking status (n=4,722)  Never smokers^b^  Past smokers^c^  Current smokers^d^  *Missing (n=267)* | 54.4 (50.5 – 58.4)  37.2 (31.6 – 42.8)  52.7 (48.4 – 57.1)  86.2 (81.0 – 91.4) | 45.6 (41.6 – 49.5)  62.8 (57.2 – 68.4)  47.3 (42.9 – 51.6)  13.8 (8.6 – 19.0) | Reference  17.3 (12.6, 21.9)  1.7 (-3.8, 7.1) | **<0.001** |
| Heavy alcohol drinking (n=3,891)  No  Yes^e^  *Missing (n=1,122)* | 41.1 (36.5 – 45.7)  52.4 (48.3 – 56.5)  52.9 (47.6 – 58.2) | 58.9 (54.3 – 63.5)  47.6 (43.9 – 51.7)  47.1 (41.8 – 52.4) | Reference  -11.3 (-14.9, -7.7) | **<0.001** |
| HbA1c, %  Mean (95%CI)  Diabetes categories^f^  Normal  Prediabetes  Diabetes | 5.4 (5.4 – 5.5)  59.9 (55.8 – 64.0)  40.3 (37.1 – 43.5)  19.1 (16.5 – 21.8) | 5.9 (5.8 – 5.9)  40.1 (36.0 – 44.2)  59.7 (56.5 – 62.9)  80.9 (78.2 – 83.5) | 0.4 (0.4, 0.5)  Reference  19.6 (15.8 – 23.4)  40.8 (37.3 – 44.3) | **<0.001**  **<0.001** |
| HIV co-infection status (n=3,389)  Negative  Positive  *Missing (n=1,600)* | 60.7 (57.3 – 64.2)  78.4 (54.8 – 100.0)  25.3 (21.7 – 28.9) | 39.3 (35.8 – 42.7)  21.6 (0 – 45.2)  74.7 (71.1 – 78.3) | Reference  -17.7 (-43.6, 8.3) | 0.222 |
| ***Dyslipidemia Measures*** |  |  |  |  |
| HDL (mg/dL) (n=4,889)  Mean (95%CI)  HDL levels^g^  Normal  Lower  *Missing (n=100)* | 53.2 (52.1 – 54.3)  53.1 (48.9 – 57.3)  47.1 (43.5 – 50.7)  37.0 (25.2 – 48.7) | 52.3 (51.0 – 53.6)  46.9 (42.7 – 51.1)  52.9 (49.3 – 56.5)  63.0 (51.3 – 74.8) | -0.9 (-2.0, 0.1)  Reference  6.0 (2.4, 9.6) | 0.083  **<0.001** |
| LDL^h^ (mg/dL) (n=2,236)  Mean (95%CI)  LDL levels  Normal (<130 mg/dL)  Elevated (130 – 159 mg/dL)  High (≥160 mg/dL)  *Missing (n=67)* | 113.2 (110.5 – 115.8)  53.7 (48.5 – 58.8)  55.8 (48.7 – 62.9)  38.7 (28.1 – 49.3)  31.7 (19.9 – 43.5) | 116.4 (113.0 – 119.8)  46.3 (41.2 – 51.5)  44.2 (37.1 – 51.3)  61.3 (50.7 – 71.9)  68.3 (56.5 – 80.1) | 3.2 (-1.1, 7.6)  Reference  -2.1 (-11.3, 7.1)  15.0 (4.7, 25.3) | 0.135  **0.011** |
| Total Cholesterol (mg/dL) (n=4,889)  Mean (95%CI)  Total cholesterol levels  Low (≤130 mg/dL)  Normal (131 – 199 mg/dL)  Elevated (≥200 mg/dL)  *Missing (n=100)* | 190.3 (187.7 – 192.8)  50.7 (44.3 – 57.0)  55.3 (50.9 – 59.8)  46.4 (41.1 – 51.8)  37.0 (25.2 – 48.7) | 198.6 (194.7 – 202.4)  49.3 (43.0 – 55.7)  44.7 (40.2 – 49.1)  53.6 (48.2 – 58.9)  63.0 (51.3 – 74.8) | 8.3 (3.4, 13.2)  Reference  -4.7 (-10.6, 1.3)  4.2 (-3.3, 11.7) | **0.002**  **<0.001** |
| Triglyceride^h^ (mg/dL) (n=2,276)  Mean (95%CI)  Triglyceride levels  Optimal (<150 mg/dL)  Elevated (150 – 199 mg/dL)  High (≥200 mg/dL)  *Missing (n=27)* | 111.5 (105.4 – 117.6)  57.9 (54.2 – 61.6)  41.8 (34.1 – 49.5)  28.7 (21.8 – 35.6)  25.7 (6.7 – 44.8) | 148.8 (134.6 – 162.9)  42.1 (38.4 – 45.8)  58.2 (50.5 – 65.9)  71.3 (64.4 – 78.2)  74.3 (55.2 – 93.3) | 37.3 (26.3, 48.2)  Reference  16.1 (9.8, 22.5)  29.2 (22.4, 36.1) | **<0.001**  **<0.001** |
| Any dyslipidemia^i&h^ (n=2,277)  No  Yes  *Missing (n=26)* | 61.0 (56.7 – 65.4)  47.4 (41.9 – 52.8)  24.6 (6.0 – 43.2) | 39.0 (34.6 – 43.3)  52.6 (47.2 – 58.1)  75.4 (56.8 – 94.0) | Reference  13.7 (7.7, 19.6) | **<0.001** |
| Statin prescription^j^ (n=2,770)  No  Yes  *Missing (n=2,238)* | 44.8 (40.0 – 49.6)  20.6 (16.0 – 25.2)  69.8 (66.6 – 73.0) | 55.2 (50.4 – 60.0)  79.4 (74.8 – 84.0)  30.2 (27.0 – 33.4) | Reference  24.2 (17.6, 30.9) | **<0.001** |
| CHD^k^ (n=4,712)  No  Yes  *Missing (n=277)* | 50.9 (47.2 – 54.6)  15.3 (5.9 – 24.8)  85.6 (80.4 – 90.8) | 49.1 (45.4 – 52.8)  84.7 (75.2 – 94.1)  14.4 (9.2 – 19.6) | Reference  35.6 (25.0, 46.1) | **<0.001** |
| Heart attack^l^ (n=4,723)  No  Yes  *Missing (n=266)* | 50.8 (47.1 – 54.5)  20.9 (11.6 – 30.2)  86.1 (80.7 – 91.5) | 49.2 (45.5 – 52.9)  79.1 (69.8 – 88.4)  13.9 (8.5 – 19.3) | Reference  29.9 (18.5, 41.4) | **<0.001** |
| Stroke^m^ (n=4,725)  No  Yes  *Missing (n=264)* | 50.9 (47.3 – 54.4)  15.6 (8.8 – 22.4)  86.9 (82.1 – 91.6) | 49.1 (45.6 – 52.7)  84.4 (77.6 – 91.2)  13.1 (8.4 – 17.9) | Reference  35.3 (28.1, 42.5) | **<0.001** |
| Abbreviations:  BMI – body mass index; CI – confidence interval; HDL – high-density lipoprotein; LDL – low-density lipoprotein; NHANES – National Health and Nutrition Examination Survey; QFT - QuantiFERON Gold-In-Tube; TST – tuberculin skin test  ^*^Mean/prevalence difference was calculated by setting those without TBI (i.e., QFT negative) as the referent group, unless indicated otherwise (with “reference” statement)  ^†^P-values from Rao-Scott Chi-square tests, unless indicated otherwise  ^‡^P-values from t-tests  ^a^Systolic ≥130mmHg and/or diastolic ≥80mmHg or any previous diagnosis of high blood pressure by health providers  ^b^Survey participants answered “No” to the question “(Have you/has SP) smoked at least 100 cigarettes in life?  ^c^Survey participants answered “Not at all” to the question “(Do you/does SP) now smoke cigarettes?” and “Yes” to the question “(Have you/has SP) smoked at least 100 cigarettes in life?  ^d^Survey participants answered “Every day” or “Some days” to the question “(Do you/does SP) now smoke cigarettes?” and “Yes” to the question “(Have you/has SP) smoked at least 100 cigarettes in life?  ^e^Survey participants answered “Yes” to the question “Was there ever time or times in (your/SP’s) life when (you/he/she) drank 4 (for female) or 5 (for male) or more drinks of any kind of alcoholic beverage almost every day?”  ^f^Diabetes was categorized according to HbA1c levels and self-reported previous type-2 diabetes mellitus diagnosis by health care providers  ^g^HDL level was using gender-specific cut-offs: “normal” HDL was defined if HDL level was ≥40 mg/dL for male or ≥50 mg/dL for female; and “lower” HDL was defined if HDL level was <40 mg/dL for male or <50 mg/dL for female  ^h^LDL and triglyceride measurements were done among a subset of survey participants who were fasting and appropriate weight variable (for those who were fasting) was applied accordingly  ^i^Any dyslipidemia was defined as having either elevated LDL, total cholesterol, triglyceride, or lower HDL levels  ^j^Taken statin in the past 30 days prior to survey date, survey participants were also asked to show medicine container to surveyor/enumerator  ^k^Survey participants answered “Yes” to the question “Has a doctor or other health professional ever told (you/SP) that (you/s/he) had coronary heart disease?”  ^l^Survey participants answered “Yes” to the question “Has a doctor or other health professional ever told (you/SP) that (you/s/he) had a heart attack (also called myocardial infarction)?”  ^m^Survey participants answered “Yes” to the question “Has a doctor or other health professional ever told (you/SP) that (you/s/he) had a stroke?”  **Bold** indicates that the finding is statistically significant at α=0.05 | | | | |

**Table S3.** Weighted prevalence of various hypertension classifications by interferon gamma tuberculosis antigen responses among representative of civilian, non-institutionalized US adult population, NHANES 2011-2012

| **Hypertension Measures** | **Weighted Prevalence (95%CI)** | | | **Prevalence Difference (95%CI)** | | |
| --- | --- | --- | --- | --- | --- | --- |
|  | **QFT Negative**  **N=4510**  **94.3% (93.3 – 95.2)** | **QFT Positive** | |  |  |  |
|  |  | **Ag-NIL Values^*^** | |  |  |  |
|  |  | **Low (<4 IU/ml)**  **N=299**  **4.0% (3.2 – 4.7)** | **High (≥4 IU/ml)**  **N=180**  **1.7% (1.2 – 2.3)** | **Low Ag-NIL vs. QFT (-)** | **High Ag-NIL vs. QFT (-)** | **High vs. Low Ag-NIL** |
| **Primary study outcome** |  |  |  |  |  |  |
| Any hypertension indication^a^ | 48.3 (44.5, 52.1) | 57.6 (48.7, 66.6) | 60.4 (53.0, 67.7) | 9.4 (1.6, 17.1) | 12.1 (3.6, 20.5) | 2.7 (-10.1, 15.5) |
| **Measured blood pressure categories** |  |  |  |  |  |  |
| Normal blood pressure^b^ | 47.9 (44.6, 51.2) | 35.6 (25.1, 46.1) | 39.5 (29.3, 49.7) | -12.3 (-22.7, -1.9) | -8.4 (-18.1, 1.2) | 3.8 (-9.7, 17.4) |
| Borderline hypertension^c^ | 17.6 (15.9, 19.3) | 21.1 (14.2, 27.9) | 17.7 (10.3, 25.1) | 3.4 (-3.0, 9.9) | 0.1 (-7.5, 7.6) | -3.4 (-13.9, 7.2 |
| Hypertension^d^ | 34.5 (31.8, 37.2) | 43.3 (34.0, 52.7) | 42.8 (33.6, 52.1) | 8.8 (-0.4, 18.1) | 8.4 (-1.4, 18.2) | -0.5 (-14.6, 13.7) |
| Stage 1 hypertension^e^ | 24.2 (21.9, 26.5) | 28.8 (18.9, 38.8) | 33.2 (22.3, 414.1) | 4.6 (-5.7, 14.9) | 9.0(-2.7, 20.7) | 4.4 (-10.2, 19.0) |
| Stage 2 hypertension^f^ | 10.3 (8.9, 11.7) | 14.5 (10.3, 18.7) | 9.6 (5.1,14.2) | 4.2 (-0.3, 8.7) | -0.6 (-5.2, 3.9) | -4.9 (-9.0, -0.7) |
| **Hypertension Diagnosis** |  |  |  |  |  |  |
| Previously diagnosed hypertension^g^ | 30.3 (27.1, 33.6) | 35.8 (28.3, 43.3) | 44.2 (36.2, 52.2) | 5.4 (-2.5, 13.4) | 13.9 (5.0, 22.7) | 8.4 (-4.7, 21.6) |
| Self-reported current use of anti-hypertension medication^h^ | 86.3 (82.7, 90.0) | 95.0 (90.7, 98.9) | 94.4 (87.7, 100.0) | 8.5 (2.3, 14.6) | 8.1 (-0.6, 16.8) | -0.6 (-7.8, 6.9) |
| Undiagnosed hypertension^i^ | 18.0 (15.8, 20.2) | 21.9 (13.6, 30.3) | 16.2 (12.1, 20.3) | 3.9 (-4.8, 12.7) | -1.8 (-7.1, 3.4) | -5.8 (-12.7, 4.8) |
| **Hypertension Control^†^** |  |  |  |  |  |  |
| Controlled hypertension without medications^j^ | 11.8 (9.5, 14.0) | 3.5 (1.2, 5.7) | 8.3 (0.0, 17.0) | -8.3 (-11.4, -5.2) | -3.5 (-12.8, 5.8) | 4.8 (-4.7, 14.3) |
| Controlled hypertension with medications^k^ | 33.9 (28.8, 39.0) | 36.6 (25.1, 48.2) | 31.4 (17.9, 44.9) | 2.9 (-10.4, 15.9) | -2.5 (-15.1, 10.2) | -5.2 (-22.5, 12.1) |
| Uncontrolled hypertension without medications^l^ | 15.2 (12.0, 18.5) | 7.3 (2.6, 12.0) | 21.3 (7.9, 34.7) | -8.0 (-13.6, -2.3) | 6.1 (-8.3, 20.4) | 14.0 (-0.9, 27.2) |
| Uncontrolled hypertension with medications^m^ | 39.1 (35.7, 42.6) | 52.6 (40.8, 64.4) | 39.0 (30.7, 47.3) | 13.5 (-0.2, 27.1) | -0.1 (-9.5, 9.2) | -13.6 (-28.9, 1.7) |
| **Abbreviations:**  CI – confidence interval; IFN-γ - interferon gamma; QFT – QuantiFERON-TB Gold In-Tube  **^*^**Estimated by subtracting TB antigen value by TB Nil control value (LBXTBA - TBXTBN)  ^†^Calculated among those with a previous diagnosis of hypertension by healthcare providers (n=1,711)  ^a^Systolic ≥130mmHg and/or diastolic ≥80mmHg or any previous diagnosis of high blood pressure by health providers  ^b^Systolic <120 mmHg and diastolic <80 mmHg  ^c^Systolic 120-129 mmHg and diastolic <80 mmHg  ^d^Including stage 1 and 2 hypertensions (i.e., Systolic ≥130mmHg or diastolic ≥80mmHg)  ^e^Systolic 130-139 mmHg or diastolic 80-89 mmHg  ^f^Systolic ≥140 mmHg or diastolic ≥90 mmHg  ^g^Survey participants answered “yes” to the question “(Have you/has SP) ever been told by a doctor or other health professional that (you/s/he) had hypertension, also called high blood pressure?”  ^h^Among those who answered “yes” to “Because of (your/SP’s) (high blood pressure/hypertension), (have you, has s/he) ever been told to take prescribed medicine?”, survey participants also answered “yes” to the question “(Are you/Is SP) now taking prescribed medicine (for high blood pressure/hypertension)?”  ^i^Elevated blood pressure levels (Systolic ≥130mmHg or diastolic ≥80mmHg) with no prior diagnosis of hypertension by health care providers  ^j^Having systolic blood pressure <130 mmHg and a diastolic blood pressure <80 mmHg without a record of taking medications to lower blood pressure levels  ^k^Having systolic blood pressure <130 mmHg and a diastolic blood pressure <80 mmHg with a record of taking medications to lower blood pressure levels  ^l^Having systolic blood pressure ≥130 mmHg or diastolic blood pressure ≥80 mmHg without a record of taking medications to lower blood pressure levels  ^m^Having systolic blood pressure ≥130 mmHg or diastolic blood pressure ≥80 mmHg with a record of taking medications to lower blood pressure levels  **Bold** indicates that the finding is significant at α=0.05 | | | | | | |

**Table S4.** Crude and adjusted associations between interferon gamma tuberculosis antigen responses and hypertension among representative of civilian, non-institutionalized US adult population, NHANES 2011 – 2012

| **Stratification Variables** | **Prevalence Ratio (95%CI)** | | | | | |
| --- | --- | --- | --- | --- | --- | --- |
|  | **Unadjusted Estimates** | | | **Adjusted Estimates^*^** | | |
|  | **Low Ag-NIL vs. QFT (-)** | **High Ag-NIL vs. QFT (-)** | **High vs. Low Ag-NIL** | **Low Ag-NIL vs. QFT (-)** | **High Ag-NIL vs. QFT (-)** | **High vs. Low Ag-NIL** |
| **Primary study outcome** |  |  |  |  |  |  |
| Any hypertension indication^a^ | **1.19 (1.04 – 1.36)** | **1.25 (1.08 – 1.45)** | 1.05 (0.84 – 1.30) | 0.99 (0.86 – 1.15) | 1.04 (0.93 – 1.16) | 1.05 (0.85 – 1.29) |
| **Measured blood pressure categories** |  |  |  |  |  |  |
| Normal blood pressure^b^ | **0.74 (0.56 – 0.99)** | 0.82 (0.64 – 1.05) | 1.11 (0.77 – 1.59) | 0.89 (0.66 – 1.21) | 0.99 (0.80 – 1.24) | 1.12 (0.76 – 1.63) |
| Borderline hypertension^c^ | 1.20 (0.88 – 1.62) | 1.00 (0.66 – 1.54) | 0.84 (0.48 – 1.46) | 1.12 (0.82 – 1.54) | 0.94 (0.61 – 1.45) | 0.84 (0.47 – 1.50) |
| Hypertension^d^ | **1.26 (1.01 – 1.56)** | 1.24 (0.98 – 1.57) | 0.99 (0.71 – 1.37) | 1.05 (0.84 – 1.32) | 1.04 (0.87 – 1.24) | 0.99 (0.72 – 1.34) |
| Stage 1 hypertension^e^ | 1.19 (0.83 – 1.77) | 1.37 (0.96 – 1.97) | 1.15 (0.72 – 1.85) | 1.06 (0.73 – 1.55) | 1.22 (0.89 – 1.66) | 1.15 (0.72 – 1.82) |
| Stage 2 hypertension^f^ | **1.41 (1.02 – 1.95)** | 0.94 (0.59 – 1.50) | **0.67 (0.45 – 0.99)** | 1.03 (0.74 – 1.44) | 0.70 (0.40 – 1.23) | 0.67 (0.43 – 1.05) |
| **Hypertension Diagnosis** |  |  |  |  |  |  |
| Previously diagnosed hypertension^g^ | 1.18 (0.94 – 1.48) | **1.46 (1.17 – 1.81)** | 1.24 (0.89 – 1.72) | 0.93 (0.74 – 1.16) | 1.16 (0.97 – 1.38) | 1.25 (0.93 – 1.66) |
| Self-reported current use of anti-hypertension medication^h^ | **1.10 (1.03 – 1.18)** | 1.09 (0.99 – 1.20) | 1.00 (0.92 – 1.08) | **1.07 (1.00 – 1.14)** | 1.07 (0.98 – 1.17) | 1.00 (0.92 – 1.09) |
| Undiagnosed hypertension^i^ | 1.22 (0.81 – 1.83) | 0.90 (0.66 – 1.23) | 0.74 (0.47 – 1.17) | 1.12 (0.73 – 1.70) | 0.82 (0.60 – 1.12) | 0.73 (0.46 – 1.17) |
| **Hypertension Control^†^** |  |  |  |  |  |  |
| Controlled hypertension without medications^j^ | **0.30 (0.15 – 0.58)** | 0.70 (0.23 – 2.12) | 2.37 (0.62 – 9.12) | **0.53 (0.28 – 1.00)** | 0.97 (0.39 – 2.39) | 1.83 (0.62 – 5.38) |
| Controlled hypertension with medications^k^ | 1.08 (0.75 – 1.55) | 0.93 (0.62 – 1.39) | 0.86 (0.51 – 1.44) | 1.01 (0.70 – 1.46) | 0.89 (0.59 – 1.35) | 0.88 (0.52 – 1.50) |
| Uncontrolled hypertension without medications^l^ | **0.48 (0.24 – 0.94)** | 1.40 (0.70 – 2.81) | **2.93 (1.34 – 6.40)** | 0.61 (0.31 – 1.21) | 1.56 (0.70 – 3.47) | **2.57 (1.14 – 5.76)** |
| Uncontrolled hypertension with medications^m^ | **1.34 (1.02 – 1.77)** | 1.00 (0.79 – 1.27) | 0.74 (0.53 – 1.03) | 1.22 (0.92 – 1.63) | 0.98 (0.75 – 1.27) | 0.80 (0.57 – 1.11) |
| **Abbreviations:**  CI – confidence interval; QFT – QuantiFERON-TB Gold In-Tube  ^*^Models adjusted for age (continuous) and gender  ^†^Calculated among those with a previous diagnosis of hypertension by healthcare providers (n=1,711)  ^a^Systolic ≥130mmHg and/or diastolic ≥80mmHg or any previous diagnosis of high blood pressure by health providers  ^b^Systolic <120 mmHg and diastolic <80 mmHg  ^c^Systolic 120-129 mmHg and diastolic <80 mmHg  ^d^Including stage 1 and 2 hypertensions (i.e., Systolic ≥130mmHg or diastolic ≥80mmHg)  ^e^Systolic 130-139 mmHg or diastolic 80-89 mmHg  ^f^Systolic ≥140 mmHg or diastolic ≥90 mmHg  ^g^Survey participants answered “yes” to the question “(Have you/has SP) ever been told by a doctor or other health professional that (you/s/he) had hypertension, also called high blood pressure?”  ^h^Among those who answered “yes” to “Because of (your/SP’s) (high blood pressure/hypertension), (have you, has s/he) ever been told to take prescribed medicine?”, survey participants also answered “yes” to the question “(Are you/Is SP) now taking prescribed medicine (for high blood pressure/hypertension)?”  ^i^Elevated blood pressure levels (Systolic ≥130mmHg or diastolic ≥80mmHg) with no prior diagnosis of hypertension by health care providers  ^j^Having systolic blood pressure <130 mmHg and a diastolic blood pressure <80 mmHg without a record of taking medications to lower blood pressure levels  ^k^Having systolic blood pressure <130 mmHg and a diastolic blood pressure <80 mmHg with a record of taking medications to lower blood pressure levels  ^l^Having systolic blood pressure ≥130 mmHg or diastolic blood pressure ≥80 mmHg without a record of taking medications to lower blood pressure levels  ^m^Having systolic blood pressure ≥130 mmHg or diastolic blood pressure ≥80 mmHg with a record of taking medications to lower blood pressure levels  **Bold** indicates that the finding is significant at α=0.05 | | | | | | |

**Table S5.** The crude and adjusted prevalence odds ratios of any hypertension stratified by race, body mass index category, and foreign-born status, among representative of civilian, non-institutionalized US adult population, NHANES 2011 – 2012

| **Stratification Variables** | **QFT Status** | **Unweighted frequency**  **Hypertension*/Total** | **Weighted Prevalence of Hypertension* (95%CI)** | **Prevalence Difference**  **(95%CI)** | **Prevalence Ratios** | |
| --- | --- | --- | --- | --- | --- | --- |
|  |  |  |  |  | **Crude** | **Adjusted**^†^ |
|  |  |  |  |  | **cPR (95%CI)** | **aPR (95%CI)** |
| All cohort | Negative  Positive | 2294/4510  286/479 | 48.3 (44.5 – 52.1)  58.5 (52.4 – 64.5) | Reference  10.2 (5.0, 15.4) | Reference  **1.21 (1.10 – 1.33)** | Reference  1.01 (0.92 – 1.10) |
| ***Stratified by age quartiles***^‡^ |  |  |  |  |  |  |
| Quartile 1 (18 – 31) | Negative  Positive | 253/1256  6/40 | 19.2 (16.9 – 21.5)  18.2 (0 – 37.5) | Reference  -1.0 (-20.7, 18.7) | Reference  0.95 (0.32 – 2.81) | Reference  0.81 (0.25 – 2.64) |
| Quartile 2 (32 – 47) | Negative  Positive | 512/1186  32/94 | 42.8 (37.8 – 47.8)  36.7 (20.0 – 52.4) | Reference  -6.1 (-19.6, 7.5) | Reference  0.86 (0.59 – 1.25) | Reference  0.87 (0.59 – 1.26) |
| Quartile 3 (48 – 62) | Negative  Positive | 678/1033  105/166 | 61.5 (58.0 – 65.0)  67.5 (55.0 – 80.1) | Reference  6.0 (-5.9, 18.0) | Reference  1.10 (0.92 – 1.31) | Reference  1.03 (0.88 – 1.21) |
| Quartile 4 (>62) | Negative  Positive | 851/1035  143/179 | 76.7 (72.3 – 81.1)  80.2 (72.9 – 87.5) | Reference  3.5 (-5.1, 12.1) | Reference  1.05 (0.94 – 1.17) | Reference  1.03 (0.91 – 1.17) |
| ***Stratified by age group*** |  |  |  |  |  |  |
| 18 – 49 | Negative  Positive | 1568/3454  175/307 | 42.5 (38.5 – 46.4)  51.1 (43.4 – 58.9) | Reference  8.7 (2.5, 14.8) | Reference  **1.20 (1.07 – 1.36)** | Reference  0.95 (0.84 – 1.08) |
| 50+ | Negative  Positive | 726/1056  111/172 | 65.2 (61.2 – 69.2)  72.7 (61.4 – 84.0) | Reference  7.5 (-3.1, 18.1) | Reference  1.11 (0.96 – 1.29) | Reference  1.07 (0.93 – 1.24) |
| ***Stratified by race*** | |  |  |  |  |  |
| Hispanic | Negative  Positive | 374/864  67/158 | 36.9 (31.4 – 42.5)  51.2 (42.0 – 60.4) | Reference  14.3 (7.3, 21.3) | Reference  **1.39 (1.20 – 1.60)** | Reference  0.98 (0.86 – 1.11) |
| Non-Hispanic White | Negative  Positive | 947/1769  47/71 | 49.9 (45.0 – 54.8)  67.0 (55.3 – 78.7) | Reference  17.1 (5.7, 28.6) | Reference  **1.34 (1.12 – 1.60)** | Reference  1.08 (0.91 – 1.27) |
| Non-Hispanic Black | Negative  Positive | 711/1196  80/115 | 55.7 (51.8 – 59.7)  64.6 (52.0 – 77.2) | Reference  8.9 (-3.9, 21.6) | Reference  1.16 (0.95 – 1.42) | Reference  0.86 (0.71 – 1.05) |
| Other Race/Ethnicity | Negative  Positive | 262/681  68/135 | 42.7 (37.5 – 47.9)  48.4 (41.3 – 55.6) | Reference  5.7 (-1.0, 12.4) | Reference  1.13 (0.98 – 1.31) | Reference  0.88 (0.71 – 1.09) |
| ***Stratified by body mass index category*** | | | | |  |  |
| Underweight (BMI <18.5 kg/m^2^) | Negative  Positive | 28/96  7/11 | 29.9 (22.4 – 37.5)  50.9 (10.6 – 91.2) | Reference  21.0 (-20.5, 62.6) | Reference  1.70 (0.71 – 4.05) | Reference  0.71 (0.34 – 1.51) |
| Normal (BMI 18.5 – 24.9 kg/m^2^) | Negative  Positive | 478/1367  75/149 | 31.5 (26.9 – 36.1)  49.2 (36.8 – 61.7) | Reference  17.7 (6.3, 29.2) | Reference  1.56 (1.23 – 1.98) | Reference  **1.24 (1.00 – 1.52)** |
| Overweight (BMI 25 – 29.9 kg/m^2^) | Negative  Positive | 709/1400  96/160 | 49.7 (46.2 – 53.2)  59.3 (46.0 – 72.6) | Reference  9.6 (-2.9, 22.2) | Reference  1.19 (0.97 – 1.48) | Reference  0.98 (0.81 – 1.20) |
| Obese (BMI ≥30 kg/m^2^) | Negative  Positive | 1040/1592  107/155 | 61.6 (57.2 – 66.1)  67.5 (57.9 – 77.1) | Reference  5.9 (-2.5, 14.3) | Reference  1.10 (0.97 – 1.24) | Reference  0.98 (0.89 – 1.08) |
| ***Stratified by foreign born status*** | |  |  |  |  |  |
| US Born | Negative  Positive | 1793/3341  120/172 | 50.2 (46.8 – 53.7)  65.6 (56.1 – 75.1) | Reference  15.4 (5.8, 25.0) | Reference  **1.31 (1.12 – 1.52)** | Reference  1.05 (0.92 – 1.21) |
| Foreign Born | Negative  Positive | 500/1167  166/307 | 37.7 (31.9 – 43.4)  51.3 (45.4 – 57.1) | Reference  13.6 (9.3, 17.9) | Reference  **1.36 (1.22 – 1.51)** | Reference  1.05 (0.92 – 1.21) |
| ***Stratified by current smoking status*** | | | | |  |  |
| No | Negative  Positive | 627/954  95/130 | 61.8 (56.0 – 67.7)  76.2 (66.8 – 85.6) | Reference  14.4 (4.2, 24.5) | Reference  **1.23 (1.07 – 1.42)** | Reference  1.09 (0.93 – 1.27) |
| Yes | Negative  Positive | 439/851  56/101 | 47.2 (42.5 – 52.0)  47.5 (34.4 – 60.7) | Reference  -0.3 (-14.3, 14.9) | Reference  1.01 (0.74 – 1.37) | Reference  0.89 (0.69 – 1.14) |
| ***Stratified by diabetes status*** |  |  |  |  |  |  |
| Euglycemic | Negative  Positive | 1083/2764  114/223 | 39.6 (35.4 – 43.8)  50.6 (42.6 – 58.5) | Reference  11.0 (3.0, 18.9) | Reference  **1.28 (1.08 – 1.51)** | Reference  1.01 (0.86 – 1.18) |
| Pre-diabetes | Negative  Positive | 689/1102  83/141 | 59.8 (56.6 – 63.0)  57.9 (44.2 – 71.6) | Reference  -1.9 (-15.6, 11.8) | Reference  0.97 (0.76 – 1.23) | Reference  0.95 (0.76 – 1.18) |
| Diabetes | Negative  Positive | 522/644  89/115 | 81.1 (78.3 – 83.9)  78.6 (68.7 – 88.5) | Reference  -2.5 (-13.0, 8.0) | Reference  0.97 (0.85 – 1.11) | Reference  0.94 (0.82 – 1.07) |
| ***Stratified by HIV Status*** |  |  |  |  |  |  |
| HIV negative | Negative  Positive | 1226/3130  102/243 | 39.1 (35.5 – 42.6)  43.2 (34.8 – 51.6) | Reference  4.1 (-4.3, 12.5) | Reference  1.11 (0.91 – 1.35) | Reference  0.93 (0.81 – 1.07) |
| HIV positive | Negative  Positive | 4/15  1/1 | 18.4 (0 – 39.0)  100.0 (100.0 – 100.0) | Reference  81.6 (61.0 – 100.0) | Reference  **5.43 (1.92 – 15.36)** | Reference  6.24 (1.79 – 21.72) |
| aPR – adjusted prevalence ratio; CI – Confidence interval; PR – prevalence ratio; QFT – QuantiFERON-TB Gold In-Tube; US – United States  ^*^Systolic ≥130mmHg and/or diastolic ≥80mmHg or any previous diagnosis of high blood pressure by health providers  ^†^Adjusted for age (continuous) and gender  ^‡^Adjusted for gender | | | | | | |

**Table S6.** Weighted prevalence of various hypertension classifications by confirmed tuberculosis infection status among representative of civilian, non-institutionalized US adult population, NHANES 2011-2012

| **Hypertension Measures** | **Weighted Prevalence (95%CI)** | | | | |
| --- | --- | --- | --- | --- | --- |
|  | **Confirmed TB Infection Status**  **N=4,266** | | | | |
|  | **Confirmed** | | **Discordant TST and QFT** | | |
|  | **Negative**  **N=3706**  **92.2% (90.5 – 93.9)** | **Positive**  **N=190**  **2.1% (1.4 – 2.8)** | **TST**^*^ **– and QFT +**  **N=177**  **2.5 (1.4 – 3.5)** | **TST + and QFT –**  **N=193**  **3.2 (2.5 – 4.00)** | **Any Discordance**  **N=370**  **5.7% (4.6 – 6.8)** |
| **Primary study outcome** |  |  |  |  |  |
| Any hypertension indication^a^ (n=2,250/4,266) | 49.6 (45.7 – 53.5) | 60.8 (51.4 – 70.3) | 50.5 (38.9 – 62.2) | 54.4 (43.5 – 65.4) | 52.7 (43.9 – 61.6) |
| **Measured blood pressure categories** |  |  |  |  |  |
| Normal blood pressure^b^ (n=1,914) | 47.0 (42.9 – 51.1) | 36.6 (27.6 – 45.5) | 49.8 (40.9 – 58.7) | 39.6 (26.1 – 53.0) | 44.0 (35.2 – 52.9) |
| Borderline hypertension^c^ (n=714) | 17.8 (15.5 – 20.0) | 15.3 (8.3 – 22.3) | 16.3 (8.2 – 24.4) | 25.1 (14.7 – 35.5) | 21.3 (13.4 – 29.1) |
| Hypertensiond (n=1,638/4,266) | 35.2 (32.3 – 38.1) | 48.1 (38.6 – 57.6) | 33.9 (25.4 – 42.4) | 35.3 (26.9 – 43.7) | 34.7 (28.3 – 41.1) |
| Stage 1 hypertension^e^ (n=1121) | 24.9 (22.5 – 27.3) | 37.0 (28.5 – 45.4) | 25.4 (16.7 – 34.1) | 24.0 (12.6 – 35.4) | 24.6 (16.3 – 32.9) |
| Stage 2 hypertension^f^ (n=517) | 10.3 (8.9 – 11.7) | 11.1 (6.2 – 16.1) | 8.5 (3.3 – 13.7) | 11.3 (4.0 – 18.5) | 10.1 (5.5 – 14.6) |
| **Hypertension Diagnosis** |  |  |  |  |  |
| Previously diagnosed hypertension^g^ (n=1,496/4,266) | 30.9 (27.5 – 34.3) | 35.8 (27.5 – 44.0) | 29.4 (17.9 – 40.8) | 37.1 (25.9 – 48.4) | 33.8 (27.0 – 40.6) |
| Self-reported current use of anti-hypertension medication^h^ (n=1,292/1,496) | 86.0 (82.2 – 89.9) | 90.2 (79.7 – 100.0) | 81.5 (65.8 – 97.1) | 98.6 (96.0 – 100.0) | 92.5 (87.4 – 97.5) |
| Undiagnosed hypertension^i^ (n=754/4,266) | 18.7 (16.4 – 21.0) | 25.2 (18.1 – 32.3) | 21.4 (12.2 – 30.6) | 17.3 (6.1 – 28.5) | 19.1 (12.2 – 25.9) |
| **Hypertension Control (n=1,496)** |  |  |  |  |  |
| Controlled hypertension without medications^j^ (n=1,286) | 11.8 (9.6, 13.9) | 6.9 (0.0, 15.0) | 13.5 (1.6, 25.4) | 5.4 (1.0, 9.8) | 8.4 (3.5, 13.3) |
| Controlled hypertension with medications^k^ (n=79) | 34.8 (29.2, 40.4) | 28.9 (16.2, 41.6) | 43.6 (20.8, 66.4) | 46.1 (34.0, 58.2) | 45.2 (35.4, 55.0) |
| Uncontrolled hypertension without medications^l^ (n=51) | 15.0 (11.5, 18.4) | 17.2 (5.7, 28.7) | 18.9 (6.1, 29.7) | 5.4 (0.3, 10.5) | 10.1 (5.5, 14.7) |
| Uncontrolled hypertension with medications^m^ (n=80) | 38.5 (34.7, 42.2) | 47.0 (36.2, 57.8) | 25.0 (12.3, 37.7) | 43.1 (28.9, 57.3) | 36.3 (26.4, 46.2) |
| **Abbreviations:**  CI – confidence interval; QFT – QuantiFERON-TB Gold In-Tube; TST – tuberculin skin test  ^*^TST positive was defined as skin induration ≥5mm among HIV-positive individuals or >10mm among HIV negative (following NHANES analytical notes). Induration <5mm (for HIV-positive individuals) or ≤10mm (for HIV-negative individuals) was considered negative  ^a^Systolic ≥130mmHg and/or diastolic ≥80mmHg or any previous diagnosis of high blood pressure by health providers  ^b^Systolic <120 mmHg and diastolic <80 mmHg  ^c^Systolic 120-129 mmHg and diastolic <80 mmHg  ^d^Including stage 1 and 2 hypertensions (i.e., Systolic ≥130mmHg or diastolic ≥80mmHg)  ^e^Systolic 130-139 mmHg or diastolic 80-89 mmHg  ^f^Systolic ≥140 mmHg or diastolic ≥90 mmHg  ^g^Survey participants answered “yes” to the question “(Have you/has SP) ever been told by a doctor or other health professional that (you/s/he) had hypertension, also called high blood pressure?”  ^h^Among those who answered “yes” to “Because of (your/SP’s) (high blood pressure/hypertension), (have you, has s/he) ever been told to take prescribed medicine?”, survey participants also answered “yes” to the question “(Are you/Is SP) now taking prescribed medicine (for high blood pressure/hypertension)?”  ^i^Elevated blood pressure levels (Systolic ≥130mmHg or diastolic ≥80mmHg) with no prior diagnosis of hypertension by health care providers  ^j^Having systolic blood pressure <130 mmHg and a diastolic blood pressure <80 mmHg without a record of taking medications to lower blood pressure levels  ^k^Having systolic blood pressure <130 mmHg and a diastolic blood pressure <80 mmHg with a record of taking medications to lower blood pressure levels  ^l^Having systolic blood pressure ≥130 mmHg or diastolic blood pressure ≥80 mmHg without a record of taking medications to lower blood pressure levels  ^m^Having systolic blood pressure ≥130 mmHg or diastolic blood pressure ≥80 mmHg with a record of taking medications to lower blood pressure levels  **Bold** indicates that the finding is significant at α=0.05 | | | | | |

**Table S7.** Crude and adjusted associations between confirmed tuberculosis infection status and hypertension among representative of civilian, non-institutionalized US adult population, NHANES 2011 – 2012

| **Hypertension Measures** | **Measures of Association** | | | | | | |
| --- | --- | --- | --- | --- | --- | --- | --- |
|  | **Prevalence Difference (95%CI)** | | **Prevalence Ratios (PR)** | | | | |
|  |  |  | **Crude PR (95%CI)** | | **Adjusted^*^ PR (95%CI)** | | |
|  | **Confirmed TBI vs.**  **non-TBI** | **Any Discordance**  **vs. non-TBI** | **Confirmed TBI vs.**  **non-TBI** | **Any Discordance**  **vs. non-TBI** | | **Confirmed TBI vs.**  **non-TBI** | **Any Discordance**  **vs. non-TBI** |
| **Primary study outcome** |  |  |  |  | |  |  |
| Any hypertension indication^a^ | 11.3 (1.0, 21.5) | 3.2 (-5.1 – 11.5) | **1.23 (1.03 – 1.46)** | 1.06 (0.91 – 1.25) | | 1.08 (0.90 – 1.30) | 0.98 (0.84 – 1.14) |
| **Measured blood pressure categories** |  |  |  |  | |  |  |
| Normal blood pressure^b^ | -10.5 (-19.4, -1.6) | 3.0 (-12.5, 6.4) | **0.78 (0.61 – 0.99)** | 0.94 (0.76 – 1.16) | | 0.89 (0.69 – 1.15) | 1.03 (0.84 – 1.26) |
| Borderline hypertension^c^ | -2.4 (-9.5, 4.6) | 3.5 (-4.1, 11.1) | 0.86 (0.55 – 1.36) | 1.20 (0.84 – 1.71) | | 0.82 (0.51 – 1.32) | 1.15 (0.81 – 1.63) |
| Hypertension^d^ | 12.9 (2.8, 23.0) | -0.5 (-7.1, 6.1) | **1.37 (1.10 – 1.70)** | 0.99 (0.82 – 1.19) | | 1.21 (0.98 – 1.49) | 0.91 (0.75 – 1.10) |
| Stage 1 hypertension^e^ | 12.1 (2.8, 21.5) | -0.2 (-8.5, 8.0) | **1.49 (1.14 – 1.94)** | 0.99 (0.71 – 1.38) | | **1.37 (1.06 – 1.77)** | 0.93 (0.66 – 1.32) |
| Stage 2 hypertension^f^ | 0.8 (-4.1, 5.7) | -0.3 (-5.2, 4.7) | 1.08 (0.69 – 1.68) | 0.98 (0.60 – 1.59) | | 0.88 (0.53 – 1.48) | 0.86 (0.52 – 1.42) |
| **Hypertension Diagnosis** |  |  |  |  | |  |  |
| Previously diagnosed hypertension^g^ | 4.9 (-3.0, 12.7) | 2.9 (-5.0, 10.7) | 1.16 (0.93 – 1.44) | 1.09 (0.86 – 1.38) | | 0.99 (0.77 – 1.28) | 1.00 (0.81 – 1.23) |
| Self-reported current use of anti-hypertension medication^h^ | 4.2 (-8.1, 16.5) | 6.4 (0.6, 12.3) | 1.05 (0.91 – 1.20) | **1.07 (1.01 – 1.15)** | | 1.03 (0.91 – 1.18) | **1.08 (1.01 – 1.16)** |
| Undiagnosed hypertension^i^ | 6.5 (-0.3, 13.3) | 0.4 (-6.9, 7.7) | **1.35 (1.03 – 1.77)** | 1.02 (0.70 – 1.50) | | 1.26 (0.97 – 1.64) | 0.96 (0.65 – 1.41) |
| **Hypertension Control^†^** |  |  |  |  | |  |  |
| Controlled hypertension without medications^j^ | -4.9 (-14.2, 4.4) | -3.3 (-8.6, 2.0) | 0.59 (0.16 – 2.10) | 0.72 (0.39 – 1.32) | | 0.85 (0.27 – 2.70) | 0.85 (0.48 – 1.53) |
| Controlled hypertension with medications^k^ | -5.9 (-18.6, 6.8) | 10.4 (-0.6, 21.4) | 0.83 (0.54 – 1.28) | 1.30 (1.00 – 1.69) | | 0.81 (0.53 – 1.22) | 1.26 (0.97 – 1.65) |
| Uncontrolled hypertension without medications^l^ | 2.3 (-9.7, 14.2) | -4.9 (-10.3, 5.4) | 1.15 (0.57 – 2.33) | 0.68 (0.42 – 1.10) | | 1.32 (0.68 – 2.58) | 0.70 (0.41 – 1.18) |
| Uncontrolled hypertension with medications^m^ | 8.5 (-3.4, 20.4) | -2.2 (-12.4, 8.1) | 1.22 (0.94 – 1.58) | 0.94 (0.71 – 1.25) | | 1.20 (0.91 – 1.58) | **1.27 (1.05 – 1.54)** |
| **Abbreviations:**  CI – confidence interval; PR – prevalence ratio; TBI – tuberculosis infection  ^*^Models adjusted for age and gender  ^†^Calculated among those with a previous diagnosis of hypertension by healthcare providers (n=1,496)  ^a^Systolic ≥130mmHg and/or diastolic ≥80mmHg or any previous diagnosis of high blood pressure by health providers  ^b^Systolic <120 mmHg and diastolic <80 mmHg  ^c^Systolic 120-129 mmHg and diastolic <80 mmHg  ^d^Including stage 1 and 2 hypertensions (i.e., Systolic ≥130mmHg or diastolic ≥80mmHg)  ^e^Systolic 130-139 mmHg or diastolic 80-89 mmHg  ^f^Systolic ≥140 mmHg or diastolic ≥90 mmHg  ^g^Survey participants answered “yes” to the question “(Have you/has SP) ever been told by a doctor or other health professional that (you/s/he) had hypertension, also called high blood pressure?”  ^h^Among those who answered “yes” to “Because of (your/SP’s) (high blood pressure/hypertension), (have you, has s/he) ever been told to take prescribed medicine?”, survey participants also answered “yes” to the question “(Are you/Is SP) now taking prescribed medicine (for high blood pressure/hypertension)?”  ^i^Elevated blood pressure levels (Systolic ≥130mmHg or diastolic ≥80mmHg) with no prior diagnosis of hypertension by health care providers  ^j^Having systolic blood pressure <130 mmHg and a diastolic blood pressure <80 mmHg without a record of taking medications to lower blood pressure levels  ^k^Having systolic blood pressure <130 mmHg and a diastolic blood pressure <80 mmHg with a record of taking medications to lower blood pressure levels  ^l^Having systolic blood pressure ≥130 mmHg or diastolic blood pressure ≥80 mmHg without a record of taking medications to lower blood pressure levels  ^m^Having systolic blood pressure ≥130 mmHg or diastolic blood pressure ≥80 mmHg with a record of taking medications to lower blood pressure levels  **Bold** indicates that the finding is significant at α=0.05 | | | | | | | |

**Table S8.** Sensitivity analysis to account for misclassification of covariates and different ways to handle age (confounder) included in the multivariable survey-weighted robus Poisson models to estimate the association between tuberculosis infection and hypertension among representative of civilian, non-institutionalized US adult population, NHANES 2011-2012

| **Models** | **Covariate(s) included in the model** | **QFT Result** | **Adjusted Prevalence Ratios** | |
| --- | --- | --- | --- | --- |
|  |  |  | **A (Age, continuous)** | **B (Age Group - Quartiles)** |
|  |  |  | **aPR (95%CI)** | **aPR (95%CI)** |
| Model 1 | Age | Negative  Positive | Reference  1.02 (0.93 – 1.13) | Reference  1.03 (0.93 – 1.14) |
| Model 2 | Age, sex | Negative  Positive | Reference  1.01 (0.92 – 1.10) | Reference  1.01 (0.91 – 1.13) |
| Model 3 | Age, sex, BMI | Negative  Positive | Reference  1.02 (0.92 – 1.13) | Reference  1.03 (0.93 – 1.15) |
| Model 4 | Age, sex, income to poverty ratio | Negative  Positive | Reference  1.00 (0.91 – 1.09) | Reference  1.01 (0.91 – 1.12) |
| Model 5 | Age, sex, country of birth | Negative  Positive | Reference  1.05 (0.96 – 1.14) | Reference  1.07 (0.97 – 1.19) |
| Model 6 | Age, sex, income to poverty ratio, country of birth, BMI | Negative  Positive | Reference  1.05 (0.95 – 1.17) | Reference  1.08 (0.97 – 1.21) |
| Model 7 | Age, sex, income to poverty ratio, country of birth, BMI, current smoking status | Negative  Positive | Reference  1.05 (0.93 – 1.17) | Reference  1.07 (0.93- 1.24) |
| Model 8 | Age, sex, income to poverty ratio, country of birth, BMI, current smoking status, type-2 diabetes mellitus status, HIV status | Negative  Positive | Reference  1.03 (0.99 – 1.08) | Reference  1.04 (0.99 – 1.08) |
| Model 9 | Age, sex, income to poverty ratio, country of birth, BMI, type-2 diabetes mellitus status, HIV status | Negative  Positive | Reference  1.04 (0.90 – 1.20) | Reference  1.05 (1.00 – 1.09) |
| Model 10* | Age, sex, race, education attainment level, country of birth, type-2 diabetes mellitus, BMI, smoking | Negative  Positive | Reference  1.01 (0.97 – 1.06) | Reference  1.04 (0.99 – 1.09) |
| Model 11 | Age, sex, race, education attainment level, country of birth, type-2 diabetes mellitus status, self-reported previous diagnosis of coronary heart disease, heart attack, and stroke | Negative  Positive | Reference  1.00 (0.96 – 1.05) | Reference  1.03 (0.98 – 1.08) |
| Model 12 | Age, sex, race, education attainment level, country of birth, type-2 diabetes mellitus status, self-reported previous diagnosis of coronary heart disease, heart attack, and stroke, BMI, smoking | Negative  Positive | Reference  1.01 (0.96 – 1.05) | Reference  1.04 (0.99 – 1.08) |
| Model 13 | Age, sex, race education attainment level, country of birth, type-2 diabetes mellitus status, self-reported previous diagnosis of coronary heart disease, heart attack, stroke, BMI, current smoking status, heavy alcohol consumption, any dyslipidemia, statin prescription, HIV status | Negative  Positive | Reference  1.07 (0.97 – 1.18) | Reference  1.09 (1.00 – 1.18) |
